# Genetic drift versus regional spreading dynamics of COVID-19

**DOI:** 10.1101/2020.05.08.20095448

**Authors:** R. Di Pietro, M. Basile, L. Antolini, S. Alberti

## Abstract

**Background:** Current propagation models of COVID-19 are poorly consistent with existing epidemiological data and with evidence that the SARS-CoV-2 genome is mutating, for potential aggressive evolution of the disease.

**Methods:** We challenged regional versus genetic evolution models of COVID-19 at a whole-population level, over 168,089 laboratory-confirmed SARS-CoV-2 infection cases in Italy, Spain and Scandinavia. Diffusion data in Germany, France and UK provided a validation dataset of 210,239 additional cases.

**Results:** The mean doubling time of COVID-19 cases was 6.63 days in Northern versus 5.38 days in Southern Italy. Spain extended this trend of faster diffusion in Southern Europe, with a doubling time of 4.2 days. Slower doubling times were observed in Sweden (9.4 days), Finland (10.8 days), Norway (12.95 days). COVID-19 doubling time in Germany (7.0 days), France (7.5 days) and UK (7.2 days) supported the North/South gradient model. Clusters of SARS-CoV-2 mutations upon sequential diffusion across distinct geographic areas were not found to clearly correlate with regional distribution dynamics.

**Conclusions:** Acquisition of mutations, upon SARS-CoV-2 spreading across distinct geographic areas, did not distinctly associate to enhanced virus aggressiveness, and failed to explain regional diffusion heterogeneity at early phases of the pandemic. Our findings indicate that COVID-19 transmission rates associate to a sharp North/South climate gradient, with faster spreading in Southern regions. Thus, warmer climate conditions may not limit SARS-CoV-2 infectivity. Very cold regions may be better spared by recurrent courses of SARS-CoV-2 infection.

## Introduction

Studies on early dynamics of COVID-19 [1] revealed that the epidemic doubled in size every 6.4 [2] to 7.4 [1] days, with a reproductive number (R_0_) of infectious cases from 2.2 [1] to 2.7 [2]. Later investigations followed disease spreading to Singapore [3], Germany [4], France and Finland (www.ecdc.europa.eu/en/covid-19-pandemic) [5-7]. However, major uncertainties remained on SARS-CoV-2 transmission dynamics [7]. Considerable effort across major research institutions was invested into modelling SARS-CoV-2 spreading determinants. Models were generated, which took into account, among others, global traveling, population density, demographic characteristics, age distribution, social dynamics, governmental policies, air pollution, virus infectious capacity, SARS-CoV-2 containment procedures, together with economical and healthcare factors [7-12] (10.21203/rs.3.rs-82122/v1). However, limited, if any, regional heterogeneity in COVID-19 transmission could be identified using such diffusion models. We reasoned that fundamental variables were missing from current analyses, and went on for identifying such missing factor(s).

SARS-CoV-2 was suggested to be sensitive to temperature and humidity, which may affect diffusion across diverse climate areas [13] (papers.ssrn.com/sol3/papers.cfm?abstract_id=3550308; ssrn.com/abstract=3556998; www.medrxiv.org/content/10.1101/2020.02.22.20025791v1). Accordingly, initial climate-dependent propagation models predicted a limited impact of COVID-19 in the Southern hemisphere, during seasons that were infection-prone in the Northern hemisphere (papers.ssrn.com/sol3/papers.cfm?abstract_id=3550308; ssrn.com/abstract=3556998). However, early foci of infection were detected in Australia and New Zealand (Figure 1). Outbreaks were also revealed in South America and extended to Central America and Mexico. Further infection foci were revealed in Saudi Arabia and Africa, and extended to sub-Saharan countries (Tables S1, S2), questioning simple models of climate-dependent SARS-CoV-2 transmission.

**Figure 1.**
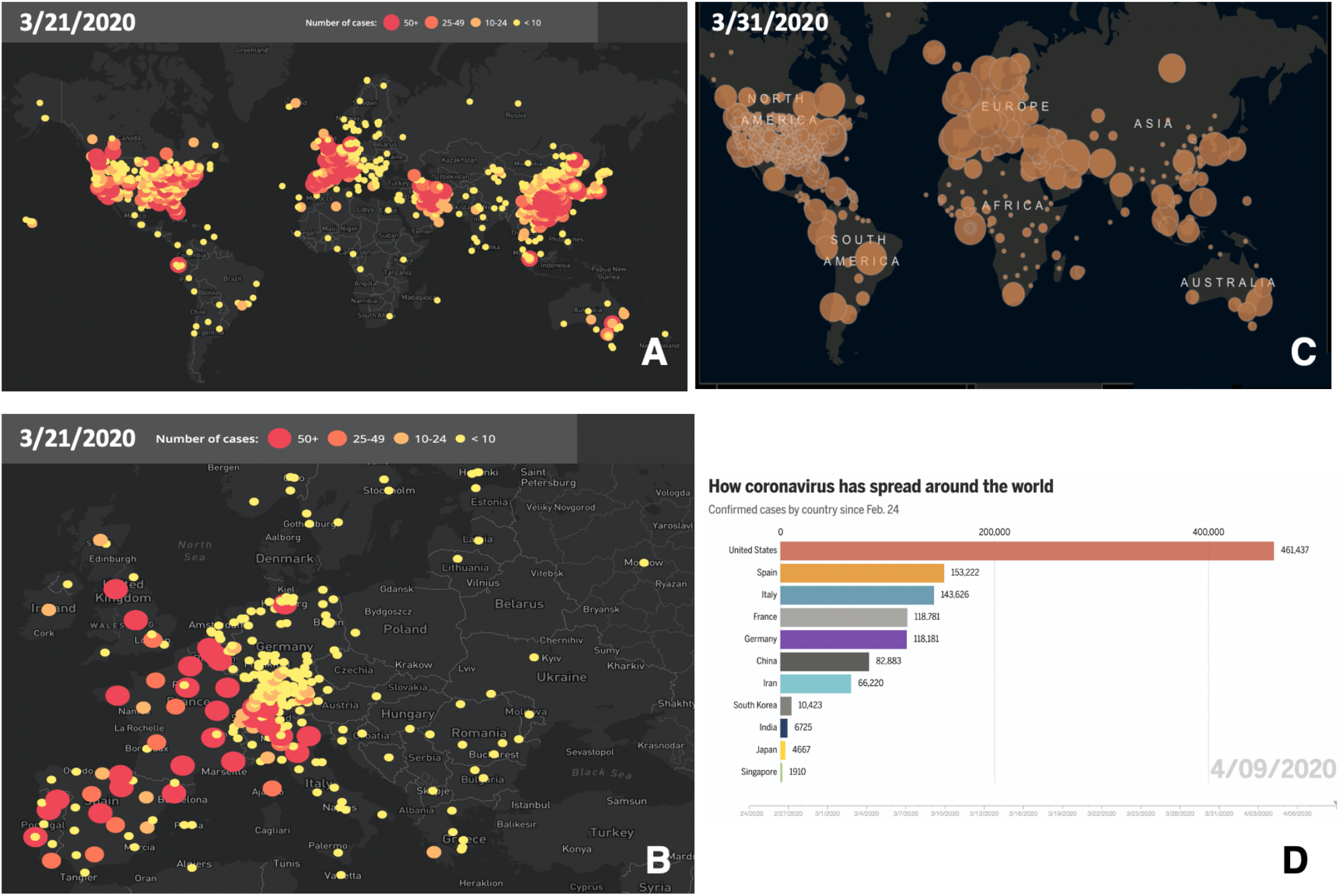
Worldwide progression of COVID-19 cases. (***A***) COVID-19 cumulative case incidence across the world, as of March 21^st^, 2020; numbers are color-coded and are proportional to circle diameter (www.healthmap.org/covid-19/). (***B***) COVID-19 cumulative case incidence, as in (***A***), zoomed over Central Europe. (***C***) COVID-19 incidence of active cases, as of March 31^st^, 2020; numbers are proportional to circle diameter (Johns Hopkins University, JHU; coronavirus.jhu.edu/map.html). (***D***) Coronavirus spreading around the world as of April 4th. Overall confirmed cases by country since February 24th (JHU, public.flourish.studio/visualisation/1694807/).

Viral evolution processes [14] may mimic regional COVID-19 spreading dynamics [15-18]. SARS-CoV-2 possesses a single-strand RNA genome [19], prone to acquire genomic mutations (nextstrain.org/ncov/; www.gisaid.org/). However, the SARS-CoV-2 RNA polymerase has error-correcting capacity, and shows replication error rates >10-fold lower than other RNA viruses [15]. Correspondingly, SARS-CoV-2 sequence diversity is low [17]. Spike proteins, in particular, show very few mutations overall [17]. Never the less, SARS-CoV-2 bearing distinct sets of mutations were isolated in different regions of the world (nextstrain.org/ncov/global), leaving the question open on whether viral genetic drift may drive evolution toward increased aggressiveness.

We thus went on to challenge regional versus genetic evolution models of COVID-19 at a population-wide level. Best chances for detecting basic transmission determinants of SARS-CoV-2 were expected before any large-scale defensive approach was implemented. Corresponding strategies were adopted to determine the impact of UV light exposure on SARS-CoV-2 diffusion [20]. Western Europe provided a vast terrain for this approach, because of the large-scale outbreaks of COVID-19 early-on in the course of the pandemic. Further advantage was provided by Europe’s high healthcare management and data collection standards (Bloomberg Global Health Index, 2018, www.bloomberg.com/; WHO, www.who.int/whr/en/; worldpopulationreview.com/countries/best-healthcare-in-the-world/; [21]), which supported a robust detection of basic diffusion parameters of COVID-19.

Broadly diverse climate regions around the CET longitude (15°E), were severely exposed to COVID-19. Spain and Italy were the countries with the highest early incidence of COVID-19 in Europe (Figures 1, S1, Table S3). The heaviest initial casualties in Italy were suffered by Lombardy and Veneto, i.e. cold and humid areas during wintertime. Markedly warmer and drier climate conditions prevail in Southern regions of the country. A further shift toward warmer/drier conditions occurs in Spain. Scandinavian countries provided a reference for cold winter temperatures, over a Sweden-Finland-Norway axis (Table S4). This offered unique opportunities to assess a climate-dependent coronavirus infection model. Such analysis was performed at a whole-population level on 86,498 laboratory-confirmed SARS-CoV-2 infection cases in Italy, 64,095 in Spain, and 17,496 cases in Scandinavia (github.com/pcm-dpc/COVID-19) (Supplemental Appendix). Diffusion data in France (Table S5), Germany (Table S6), and UK (Table S7) provided a validation dataset, encompassing 210,239 COVID-19 cases. This model was then merged with that of coronavirus genetic evolution (Figure 2), for detecting signs of positive selection for increased aggressiveness across the analyzed regions.

**Figure 2.**
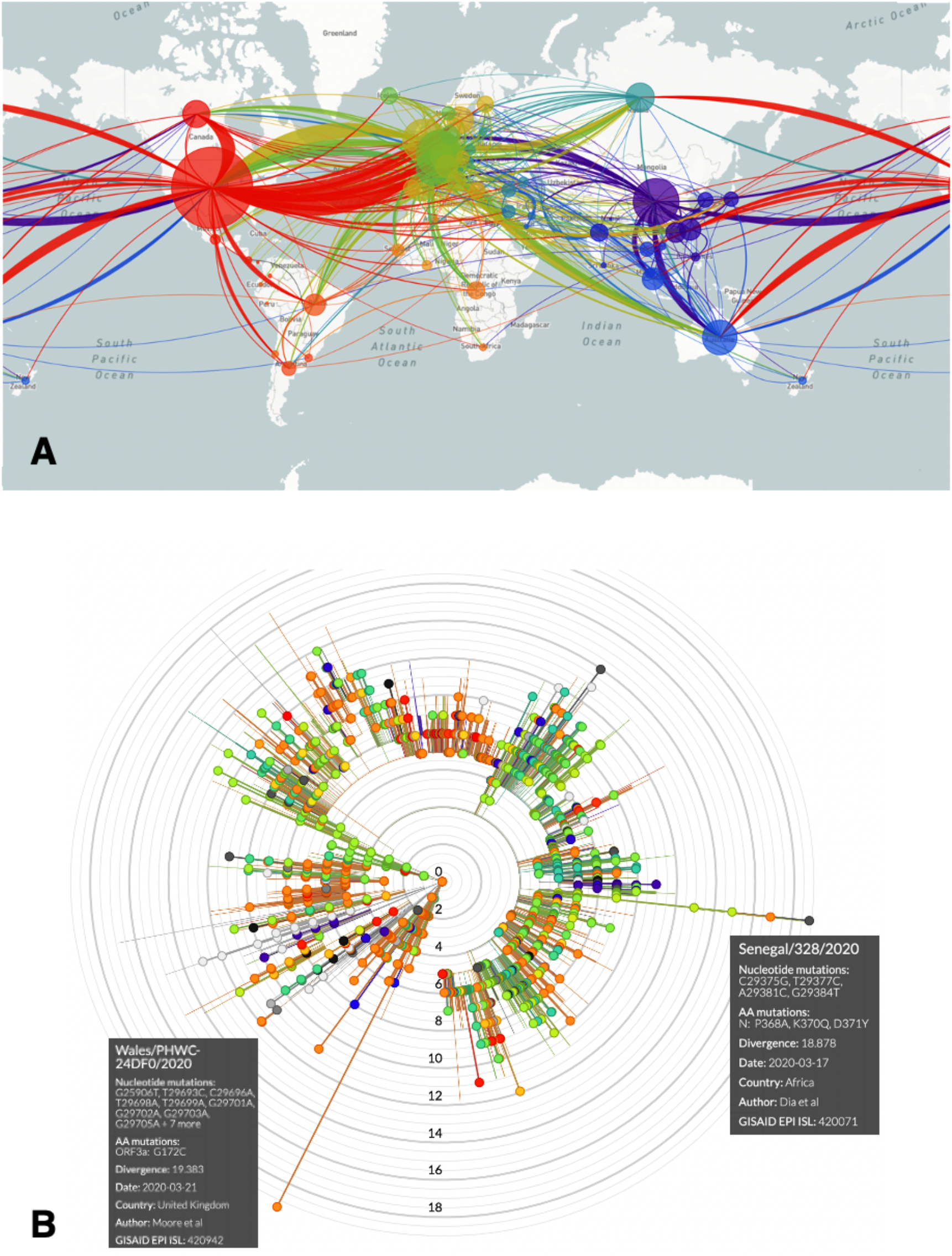
COVID-19 spreading and SARS-CoV-2 mutations. (***A***) Worldwide SARS-CoV-2 diffusion trajectories. Circle diameters are proportional to the number of virus isolates showing different sequences/acquired mutations. (***B***) Radial diagram of SARS-CoV-2 mutations worldwide. Concentric circles indicate the number of acquired genomic mutations detected in individual virus isolates.

## Methods

### Incidence data

Data on laboratory-confirmed SARS-CoV-2 infection cases in Europe were collected at early time-points of the pandemic/peak diffusion rates from the following sources: Italy (github.com/pcm-dpc/COVID-19), France (dashboard.covid19.data.gouv.fr/vue-d-ensemble?location=FRA), UK (www.nhs.uk/), Germany (<u>corona.rki.de</u>), Spain (RTVE -Ministry of Health; www.rtve.es/noticias/20200415/mapa-del-coronavirus-espana/2004681.shtml), Sweden (Public Health Agency of Sweden; www.folkhalsomyndigheten.se/smittskydd-beredskap/utbrott/aktuella-utbrott/covid-19), Finland (National Institute for Health and Welfare THL; <u>thl.fi/en/web/thlfi-en</u>), Norway (Norwegian Institute of Public Health; www.fhi.no/sv/smittsomme-sykdommer/corona/dags--og-ukerapporter/dags--og-ukerapporter-om-koronavirus).

As current transmission models <u>did not lead to predict regional heterogeneity</u> [7-12] (10.21203/rs.3.rs-82122/v1), we set our search to identify <u>additional</u> variables that <u>were not taken into account</u> by pre-existing models.

Key considerations of our strategy were:

I. An explosive diffusion of SARS-CoV-2 in Western Europe occurred early along the course of the pandemic, providing a vast number of infection cases, over largely overlapping calendar time-frames.
II. High-quality disease-reporting procedures allowed whole-population-level analyses, with inclusion of 378,328 laboratory-confirmed SARS-CoV-2 infection cases across continental Europe and UK.
III. The North-South span of the European regions involved in the early phases of COVID-19, provided vast diversity of climatic zones. The null hypothesis we challenged was that COVID-19 transmission velocity <u>would not have been different</u> across climate areas, quantitatively categorized as an independent variable.
IV. As viral evolution processes in specific geographic areas [14] can effectively mimic climate region-associated spreading [15-18], these processes were analyzed accordingly.
V. A potent confounding factor in disease transmission analyses is the founder effect, i.e. the date of first moving of infectious cases to a geographic site. To prevent this bias, the doubling number of COVID-19 cases, rather than their absolute number, was considered [20].
VI. A further confounding factor was that of large-scale virus diffusion containment measures. Hence, we directed our search toward the initial period of explosive diffusion of the virus, and ended our observations at the time of first reduction of infectious case incidence rates, upon implementation of containment measures, in a region-by-region manner [20].

Disease severity was then classified as (a) hospitalized cases, (b) intensive-care unit patients, (c) recovered cases, (d) deaths. These findings were presented as cumulative incidence by region.

The cumulative incidence of COVID-19 cases was then linked to Köppen–Geiger climate classification maps (koeppen-geiger.vu-wien.ac.at/present.htm). These were computed as mean parametrization of data collected between 1980 and 2016 [22]. The tripartite classification by country areas was compounded as an independent variable versus COVID-19 doubling time (Table 1).

**Table 1:** COVID-19 doubling time versus climate area.

| Country/region | COVID-19 doubling time (days) | Climate area | Lab-confirmed case numbers * |
| --- | --- | --- | --- |
| Spain | 4.2 | Csa/Csb/Bsk | 64,095 |
| Southern Italy | 5.38 | Csa/Csb | 5,322 |
| Central Italy | 5.87 | Csa/Cfa/Cfb | 10,842 |
| Northern Italy | 6.63 | Cfa/Cfb | 70,334 |
| Germany | 7.0 | Cfb | 73,522 |
| France | 7.5 | Cfb | 68,665 |
| UK | 7.2 | Cfb | 68,052 |
| Sweden | 9.4 | Dfc/Cfb | 11,321 |
| Finland | 10.8 | Dfc/Dfb | 2,646 |
| Norway | 12.95 | Dfc/Dfb/ET | 5,855 |

\*: Laboratory-confirmed SARS-CoV-2 infection cases in Europe cases were retrieved by country at peak diffusion rates, before the landmark dates indicated: Italy (github.com/pcm-dpc/COVID-19, March 27<sup>th</sup> 2020), France (dashboard.covid19.data.gouv.fr/vue-d-ensemble?location=FRA; April 4<sup>th</sup> 2020), UK (www.nhs.uk/; April 9<sup>th</sup> 2020), Germany (corona.rki.de; April 2<sup>nd</sup> 2020), Spain (RTVE - Ministry of Health; www.rtve.es/noticias/20200415/mapa-del-coronavirus-espana/2004681.shtml; March 27<sup>th</sup> 2020), Sweden (Public Health Agency of Sweden; www.folkhalsomyndigheten.se/smittskydd-beredskap/utbrott/aktuella-utbrott/covid-19; April 9<sup>th</sup> 2020), Finland (National Institute for Health and Welfare THL; thl.fi/en/web/thlfi-en; April 7<sup>th</sup> 2020), Norway (www.fhi.no/sv/smittsomme-sykdommer/corona/dags--og-ukerapporter/dags--og-ukerapporter-om-koronavirus; April 14<sup>th</sup> 2020).

### SARS-CoV-2 mutation analysis

SARS-CoV-2 genomic RNA sequences and country-correlated data were obtained from nextstrain.org/ncov/global. Each data-point was represented as a bead, whereby each bead corresponded to a specific set of virus mutations (mutation haplotype) (Figures 2, S2-9). ‘Beads-on-a-string’ plots were then generated, that represented linked series of individual mutation haplotypes, that acquired subsequent mutations over time. Phylogeny trees for such mutation clusters were then obtained, for drawing distinct evolutionary branches of SARS-CoV-2 (<u>nextstrain.org/ncov/europe?branchLabel=aa</u>) (Figures 2, S2-9).

### Statistical analysis

The cumulative incidence of COVID-19 cases versus calendar dates was acquired at province/county level [23, 24]. Corresponding plots acted as a smoother, for accurate determination of infection curve parameters. They also served to average urban versus countryside population dynamics/events on a province-by-province basis. Disease cumulative incidence graphs were found to largely follow a peculiar linear growth pattern [25]. This allowed to rigorously apply linear regression methodology for determining case-incidence rates.

At subsequent time points, deviations from linearity, with flattening of disease incidence curves, were recorded, following implementation of country-wide restrictions in traveling and social interactions (www.gazzettaufficiale.it/eli/gu/2020/03/08/59/sg/pdf). These inflection points were taken as landmark dates, and marked the end of the observation period. From each of these dates, the doubling time for cumulative number of diagnoses was calculated backward for each province as follows. Two dates were identified: the maximum date, at which the cumulative number of diagnoses were lower than a half of the cumulative number of diagnoses at the landmark time, and the minimum date, with a cumulative number of diagnoses greater than a half of the cumulative number of diagnoses at the landmark date. The fraction of days from the minimum date to achieve half of the cumulative number of diagnoses at the landmark date were obtained by a linear assumption for the cumulative incidence between the two dates. Correspondingly, distinct calendar dates were applied to data-collection in different provinces, regions and countries, according to the spreading sequence of the pandemic. Of note, each of these estimates corresponded to the fastest spreading velocity of COVID-19 in each region.

Coefficients, standard error, 95% confidence intervals were computed. Percentile distribution boxplots of COVID-19 cases doubling times were drawn. Median, maximum value, minimum value and distribution outliers were estimated. The correlation between COVID-19 spreading rates versus normalized climate-area values was computed by Anova.

### Software

Stata software version 16 was used for data importing, manipulation and graphics (StataCorp. 2019. *Stata Statistical Software: Release 16*. College Station, TX: StataCorp LLC).

## Results

### COVID-19 case doubling-time by geographic area

Infection transmission rates were computed for:

#### Italy

on COVID-19 cases from March 3^rd^ to March 27^th^ (n=86,498) (Supplemental Appendix) (Figures S10-12).

#### Spain

on COVID-19 cases from February 25^th^ to March 27^th^ 2020 (n=64,095) (Figure S13).

#### Norway

on data (>50 infection case outbreaks) obtained from February 21^st^ to April 14^th^ 2020 (n=6,676) (Figure S14).

#### Finland

on COVID-19 cases from March 1^st^ to April 7^th^ 2020 (n=2,646) (Figure S15).

#### Sweden

on data (>50 infection case outbreaks) obtained from February 26^th^ to April 9^th^ 2020 (n=8,995) (Figure S16).

#### France

on COVID-19 cases from February 25^th^ to April 4^th^ 2020 (Figure S17).

#### UK

on COVID-19 cases from February 1^st^ to April 9^th^ 2020 (Figure S17).

#### Germany

on COVID-19 cases from February 24^th^ to April 2^nd^ 2020 (Figure S17).

### COVID-19 doubling time versus climate region

Quantitative climate assessments are affected by complex, interdependent sets of variables [26]. Up to 89 distinct parameters are required for meteorological classification alone (apps.ecmwf.int/datasets/data/interim-full-moda/levtype=sfc/). Discrete humidity measures, temperature profiles, (papers.ssrn.com/sol3/papers.cfm?abstract_id=3556998) [13, 27] and weather structure, intertwine with lifestyle, social and occupational determinants [27] (www.medrxiv.org/content/10.1101/2020.03.23.20040501v4). Hence, fundamental sources of uncertainty are associated to climate modeling [11]. We thus resorted to utilizing the Köppen–Geiger climate classification (<u>koeppen-geiger.vu-wien.ac.at/present.htm</u>), as drawn over 30+ years of observations, and robustly validated [22, 26, 28, 29]. This was summarized as a tripartite classification of country/region/province, which was compounded as an independent variable versus COVID-19 spreading velocity (Table 1).

Cumulative numbers of COVID-19 cases versus calendar dates were normalized to the highest case incidence in each area (Figures S10-17). Pandemic doubling times were correspondingly computed (Table S3) [20] and grouped by geographic region. The average doubling time for Northern Italy was 6.63 (SD=1.94) days; 5.87 (SD=1.08) days in Central regions; 5.38 (SD=2.31) days in Southern areas, for significantly shorter doubling time in Southern regions (P=0.02 versus Northern Italy) (Table S3, Figures 3, S10-12). The mean COVID-19 doubling-time for the whole country was 6.06 (SD=1.95) days.

**Figure 3.**
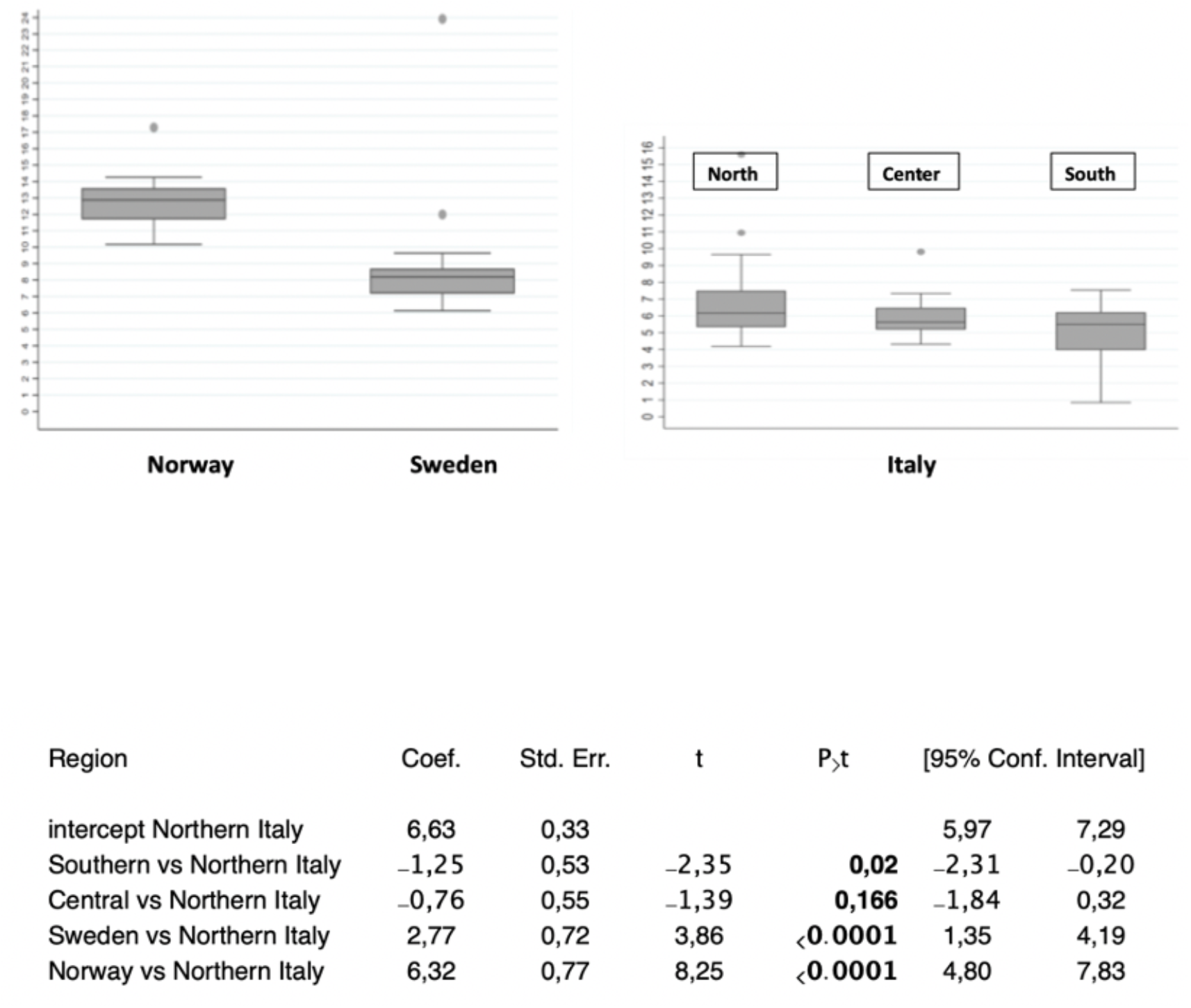
COVID-19 diffusion across geographic areas. (*top*) Distribution boxplots of SARS-CoV-2 infected cases doubling times in the areas/countries analyzed. Upper horizontal line: 75^th^ percentile; lower horizontal line: 25^th^ percentile; horizontal bar within box: median; upper horizontal bar outside box: maximum value; lower horizontal bar outside box: minimum value. Dots: distribution outliers. (*bottom*) SARS-CoV-2 infected cases doubling times versus central intercept (benchmark=Northern Italy). Coef.: coefficient; Std. Err.: standard error; 95% confidence intervals are shown. P>t: 0.002 Southern versus Northern Italy; <0.0001 Sweden versus Northern Italy; <0.0001 Norway versus Northern Italy.

With a doubling time of 4.2-days, Spain extended such a trend (Figure S13). At the opposite end of the climate spectrum, Scandinavia showed longer COVID-19 doubling times, over a Sweden-Finland-Norway axis, with a doubling time of 9.4 days (SD=1.2) for Sweden (P<0.0001 versus Northern Italy), 10.8 days for Finland, 12.95 days (SD=0.52) for Norway (P<0.0001 versus Northern Italy) (Figures 3, S14-16). This depicted a distinct North-South gradient of COVID-19 spreading velocity (Anova P<0.0001) (Figure 4, Table 1).

**Figure 4.**
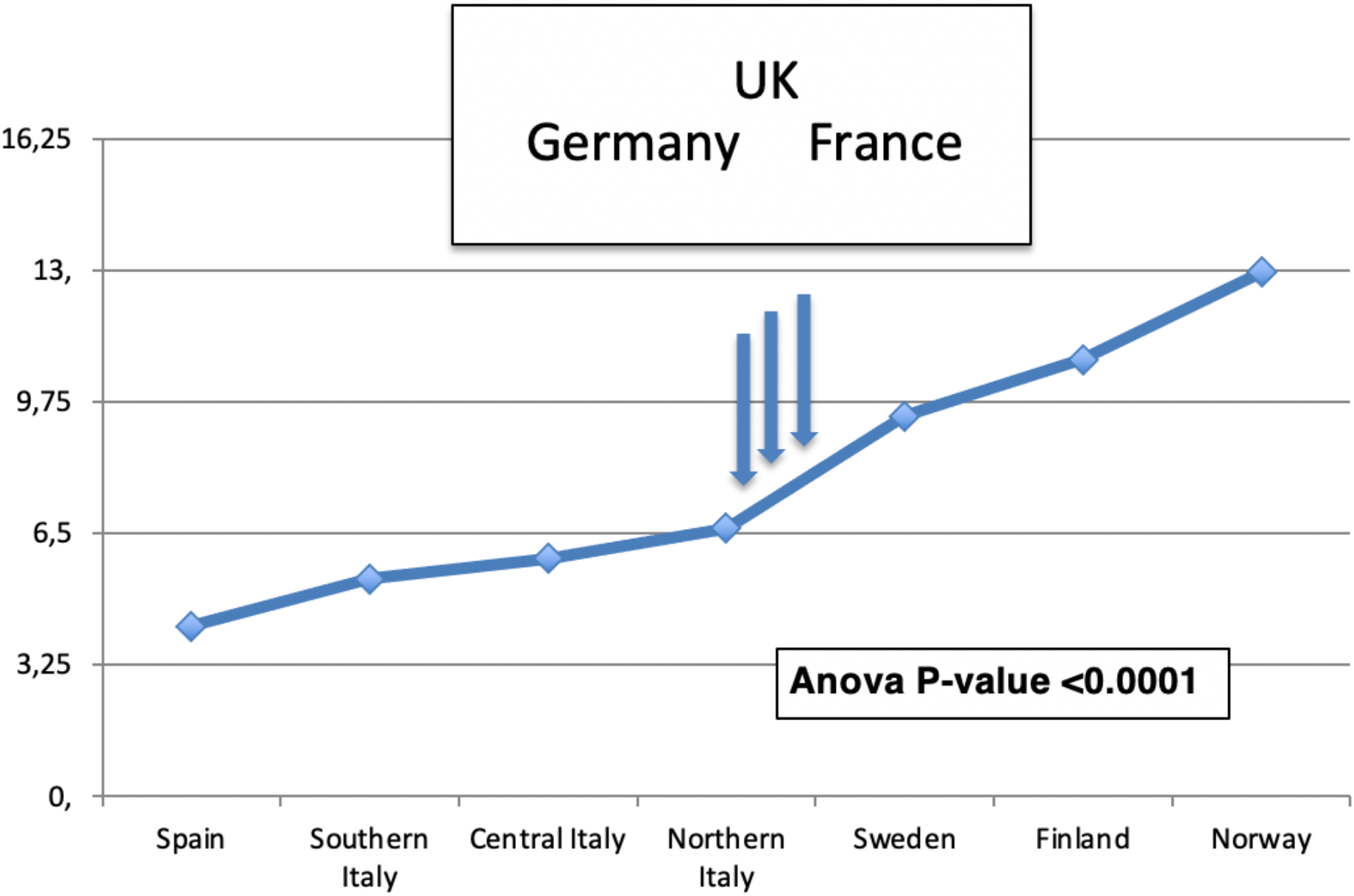
The COVID-19 North-South gradient. The North-South gradient of SARS-CoV-2 infected cases doubling times across countries is depicted. Country values were plotted according to their classification by climate zone (Table 1). The Anova P-value for Cartesian numerals association is shown. Vertical arrows: COVID-19 doubling times in validation datasets (Germany, France, UK).

This climate model was challenged versus a validation data-set of 210,239 laboratory-confirmed COVID-19 cases in Germany, France and UK. Average climate areas for all three countries fell in between classification classes of Northern Italy and Southern Sweden. Pandemic doubling time was computed to be 7.0 days in Germany (Figure S17). In sharp consistency, corresponding doubling times in France and UK were 7.5 and 7.2 days, respectively. This fell in between Northern Italy and Sweden data, as predicted by our model.

Disease severity as classified hospitalization, intensive-care unit and fatality rates were compounded as cumulative incidence by region (Figure S12). However, analysis of neither disease onset severity nor outcome provided correlation with parameters of regional diffusion heterogeneity. It should be noted that data on recoveries and deaths are not consistently classified across all the regions under study, and are considered less reliable than those of confirmed COVID-19 cases [20].

### SARS-CoV-2 genetic-drift driven diffusion

Sequence mutation analysis revealed different branches of acquired mutations, i.e. distinct groups of viral genome mutations (haplotypes), at sites of major diffusion in Europe (nextstrain.org/ncov/europe) (Figures 2, S2-9). Each of these branches was observed to acquire additional mutations over time, in an uneven manner among different geographic areas. We searched this data-set for potential indicators of positive selection for specific virus mutation(s). One virus mutation, i.e. the spike D614G amino acid change, was associated with increased COVID-19 aggressiveness [17]. The D614G mutation appeared very early in Europe (inferred date, Jan. 6, 2020) (nextstrain.org/ncov/; www.gisaid.org/) [17] and spread evenly across European countries.

Among descendants of D614G viruses, we looked for evidence of positive selection over additional mutations. It should be noted that most viral mutations may not have phenotypic effects, most of them being probably neutral or near neutral. Further, while some mutations may become dominant over time, the overall diversity of SARS-CoV-2 genomes will continue to increase due to genetic drift. Never the less, if positive selection for one or more virus mutation had been at work, deviation from even distributions of virus descendants across regions had to be expected. Among flags of such unevenness, we looked for (a) mutation-correlated increase of disease severity over time (b) prevalence of such mutation(s) in hardest-hit countries, (c) progressively broader diffusion of more aggressive virus genotypes along the course of the pandemic.

The highest numbers of accumulated mutations were revealed in SARS-CoV-2 in Wales and Senegal isolates (Figure 2B). Hence, they most likely represented late correlates of viral genetic drift over time. Of interest, the lowest number of accumulated mutations was recorded in Italy, a country with high disease severity in Europe. This appeared poorly consistent with a progressive increase of disease severity upon accumulation of novel mutations, suggesting instead correlation with a short SARS-CoV-2 evolution time. Large mutation loads were observed in Spain (n=14), the second hardest-hit country in Europe. However, a similar mutation load was observed in Sweden (n=13), a country with much more limited COVID-19 transmission and severity, further supporting correlation with genetic drift. Consistent, large mutation loads were observed in late-disease-insurgence countries, such as France and Belgium (n=16), supporting a slow SARS-CoV-2 genomic evolution, along the course of the disease (Figures S3-9).

## Discussion

Large efforts have gone into modelling COVID-19 transmission, according to global and local population dynamics, demographics, governmental policies and infectious ability of the virus [7-12] (10.21203/rs.3.rs-82122/v1). Most models, though, showed inadequate capacity for predicting regional diffusion dynamics of the pandemic [11].

We speculated that fundamental variables associated to COVID-19 uneven diffusion remained to be identified, and set a search for discovering such factor(s). We went on to perform a population-wide analysis, on 378,328 laboratory-confirmed SARS-CoV-2 infection cases in continental Europe and UK. A robust determination required collecting epidemiological data [23, 24], before intervention with disease-containment measures. We thus went on to identify landmark dates, as inflection points of disease incidence curves associated to disease-taming procedures. This led us to first identify a quasi-universal pattern of linear growth of COVID-19 cases over time, consistent with a unique diffusion mode of SARS-CoV-2 [25]. Distinct COVID-19 transmission rates were then identified as associated to different geographic regions.

Still, accumulation of mutations of SARS-CoV-2 may have led to distinct selection for disease progression over different regions. An indicator of selective pressure for viral evolution has been that of progressively larger prevalence across different geographic locations [17], as indicated for the spike D614G mutation. However, D614G was associated with higher upper respiratory tract viral loads [16-18], but not with increased disease severity. This appeared puzzling [15], as higher viral loads have been associated to worse disease course [30] and to increased mortality [31]. The hypothesis of positive selection of spike D614G was further investigated in the United Kingdom using more than 25,000 whole-genome SARS-CoV-2 sequences. This indicated 614G increases in frequency relative to 614D as consistent with selective advantage, but not in all cases [16]. This suggested that “a combination of evolutionary selection for G614 and the founder’s effects of being introduced into highly mobile populations may have together contributed in part to its rise” [17]. These findings were recapitulated “as a slow genetic drift .. of a highly stable [SARS-CoV-2] genome” [15, 32], during the analyzed time-frames.

Never the less, we looked for evidence of selection for viral evolution, utilizing broader indicators of predominance of specific mutation(s) versus disease severity. Four major mutation groups/haplotypes were revealed in all examined European countries. The highest number of accumulated mutations was revealed in Wales and Senegal SARS-CoV-2 isolates, suggesting correlation with genetic drift, at late stages of the disease. The lowest number of accumulated mutations was recorded in Italy, the country that first showed severe disease outbreaks in Europe, and similar mutation loads were observed in Spain, the second hardest-hit country in Europe, and Sweden, a country with much less explosive COVID-19 transmission, suggesting correlation with disease duration, rather than with selection for higher disease severity. The larger mutation loads were revealed in France and Belgium, both late-disease-insurgence countries, further supporting a relationship between mutation acquisition and length of disease course. Taken together, our findings add evidence to a model of SARS-CoV-2 genetic drifting during the early course of the pandemic, rather than to selective pressure for increased COVID-19 aggressiveness.

As COVID-19 spreading models based on population demographics and socio-economic factors could not account for regional diffusion heterogeneity, neither could an evolving infectious capacity of the virus, our findings supported a climate dependency of COVID-19 transmission capacity. This showed a sharp North-South gradient, with the shortest COVID-19 doubling times in Southern Italy and Spain. At the opposite end of the climate spectrum, Scandinavia showed the longest COVID-19 doubling times, over a Sweden-Finland-Norway axis. This climate model was verified in validation datasets of COVID-19 cases in Germany, France and UK, which included 210,239 laboratory-confirmed SARS-CoV-2 infection cases, and showed pandemic doubling times that were intermediate between Northern and Southern regions, in sharp consistency with the climate-area Köppen–Geiger model [22].

Findings of more efficient coronavirus spreading in warmer regions are consistent with resilience of coronaviruses to environmental conditions [33-35]. Of note, the Middle East Respiratory Syndrome (MERS) was first reported in Saudi Arabia (www.cdc.gov/coronavirus/mers). MERS is caused by the MERS-CoV, which is structurally and genetically related to SARS-CoV, indicating that at least some coronavirus strains may better propagate in high-temperature climate conditions (www.cdc.gov/coronavirus/mers/risk.html).

However, climate areas are associated to much more complex sets of variables, among them indoor versus outdoor temperature profiles, specific/relative/absolute humidity [13, 26, 27], UV exposure versus daily time/season/latitude [20, 27], weather structure and ventilation, together with social behavior, inter-individual distancing, indoor-crowding, lifestyle, outside physical activity, (papers.ssrn.com/sol3/papers.cfm?abstract_id=3556998) [27], versus community structure, socio-economical and healthcare factors (www.medrxiv.org/content/10.1101/2020.03.23.20040501v4). Recent studies have begun to dissect such determinants, indicating impact of UV exposure on COVID-19 transmission [20], while suggesting no role of temperature and humidity (SSRN 3567840, 2020 - papers.ssrn.com) [20, 29]. Our own research confirms these findings, and the lack of impact of temperature on COVID-19 progression over the areas analyzed (unpublished observations). Additional work is expected to bring-in further insight over climate area-associated virus diffusion determinants.

Taken together, our findings suggest higher SARS-CoV-2 resilience in warmer regions than previously predicted, and caution that high environmental temperatures may not efficiently tame SARS-CoV-2 infectiousness [12]. Cold regions may be better spared by recurrent courses of COVID-19.

## Data Availability

all data referred to in the manuscript are available as needed

## Acknowledgments

We are much indebted to all the information curators we cite, and to the website providers, the article data and graphic primers have been downloaded from.

## Role in the article

All authors contributed to literature search, figures, study design, data collection, data analysis, data interpretation. S.A and R.DiP. wrote the manuscript draft. All authors contributed to discussing and writing the final text. R.DiP. and M.B. contributed equally to this work.

## Footnote page

### Funding

The University of Messina provided funding to this work.

### Meeting presentation

These findings have not yet been presented to meetings.

