## Supplemental text & figures for "Genetic drift versus regional spreading dynamics of COVID-19"

### Supplemental Figures

**Figure S1. Worldwide temperature distributions and climate zones.**

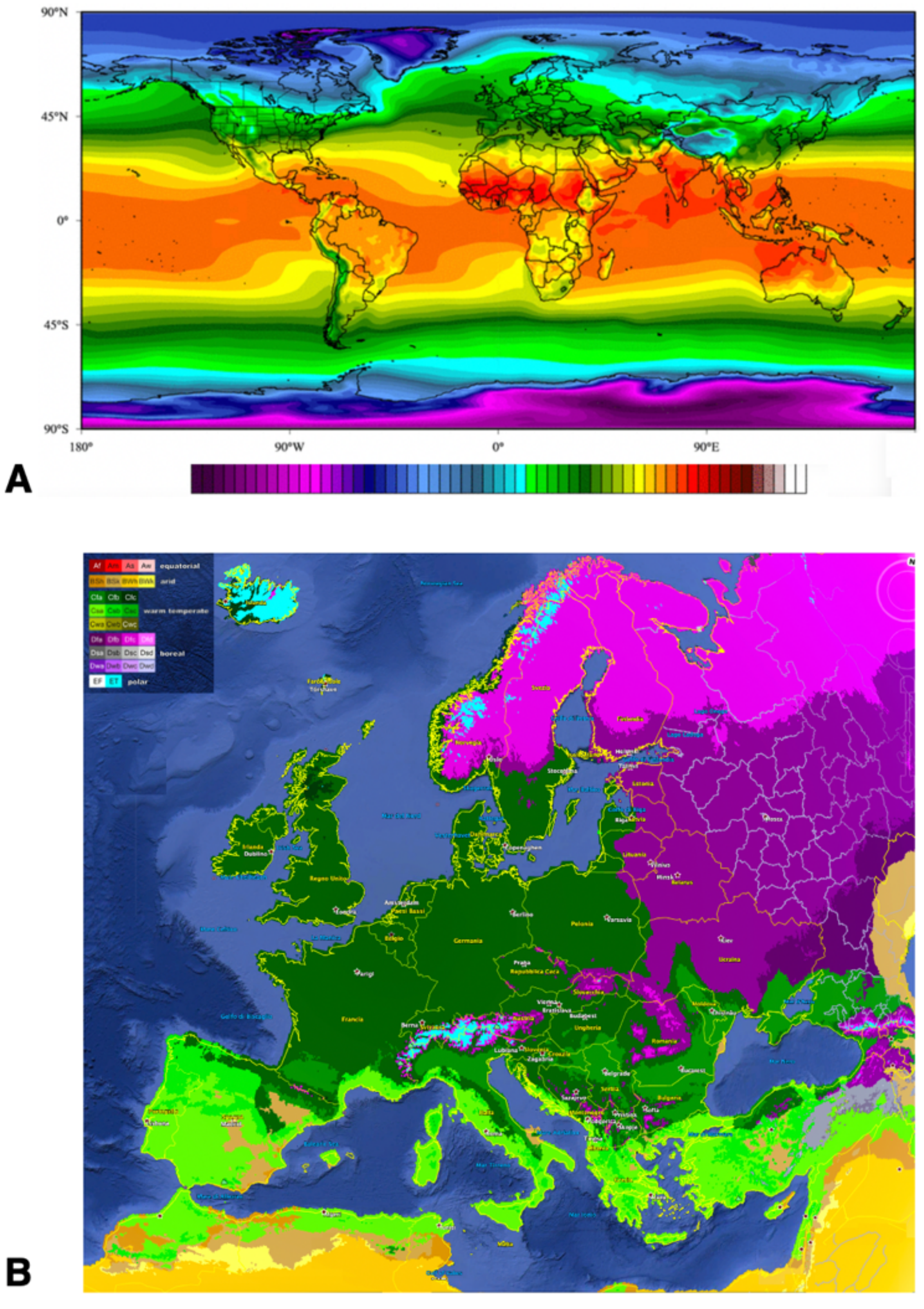

(*top*) Worldwide temperature averages (pseudocolor) versus geographic areas (average temperatures for early spring).

(*bottom*) Köppen–Geiger climate classification map of Europe.

**Figure S2. Global accumulation of genomic mutations of SARS-CoV-2.**

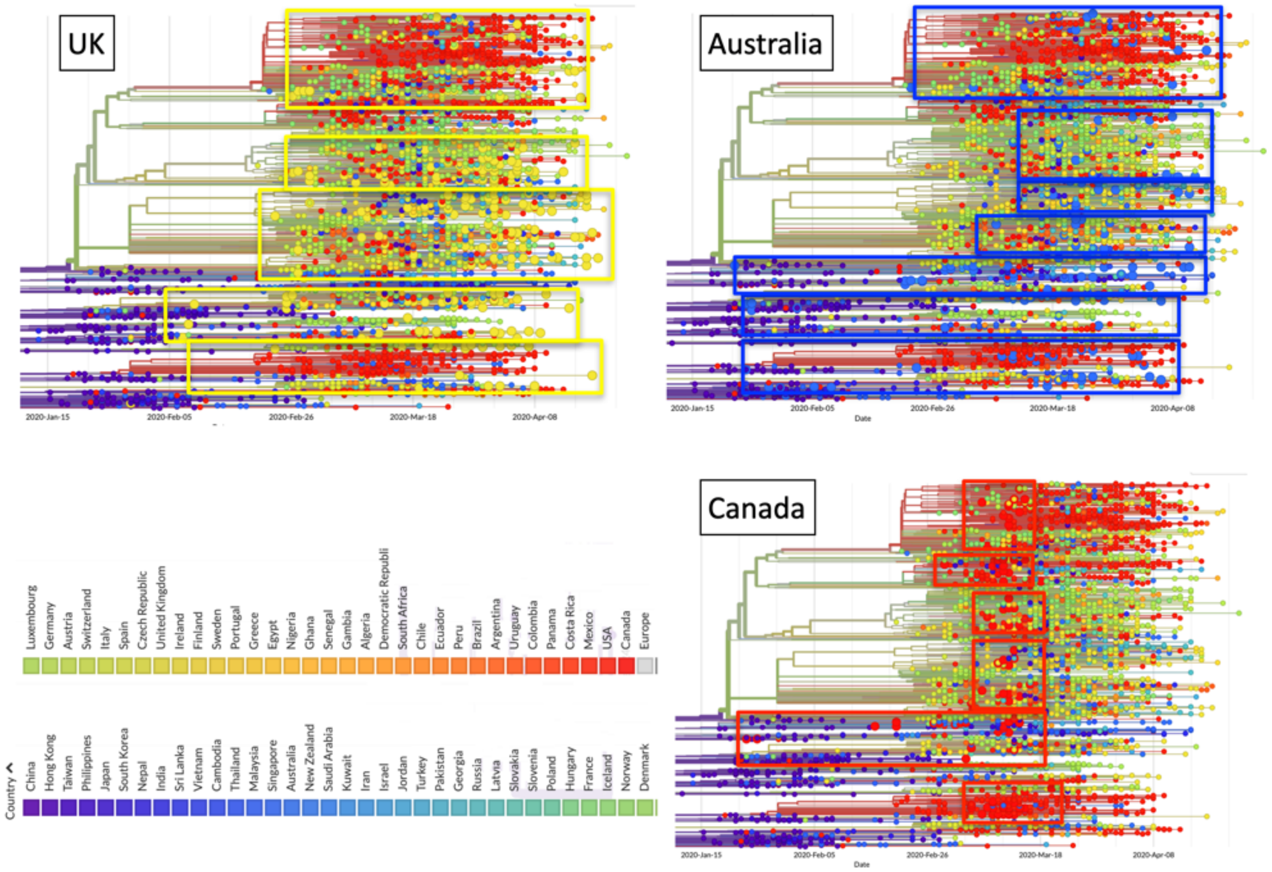

Accumulation of genomic mutations of SARS-CoV-2 over time around the globe. Each data-point is represented as a bead, whereby each bead corresponds to a specific set of virus mutations (mutation haplotype). Beads and bead sequences are color-coded, according to the country where the virus sample was isolated from (bottom left). The 'beads-on-a-string' plots link successions of viral mutations, i.e. mutation haplotypes that acquired additional mutations over time. Phylogeny trees for such mutations are presented, that draw distinct evolutionary branches of SARS-CoV-2 over calendar dates (bottom) (nextstrain.org/ncov/). Larger beads indicate mutations identified in the indicated country. UK: bright yellow; Australia: blue; Canada: red. Rectangles enclose mutations evolutionary branches by country.

**Figure S3. Genomic mutations of SARS-CoV-2 in Europe.**

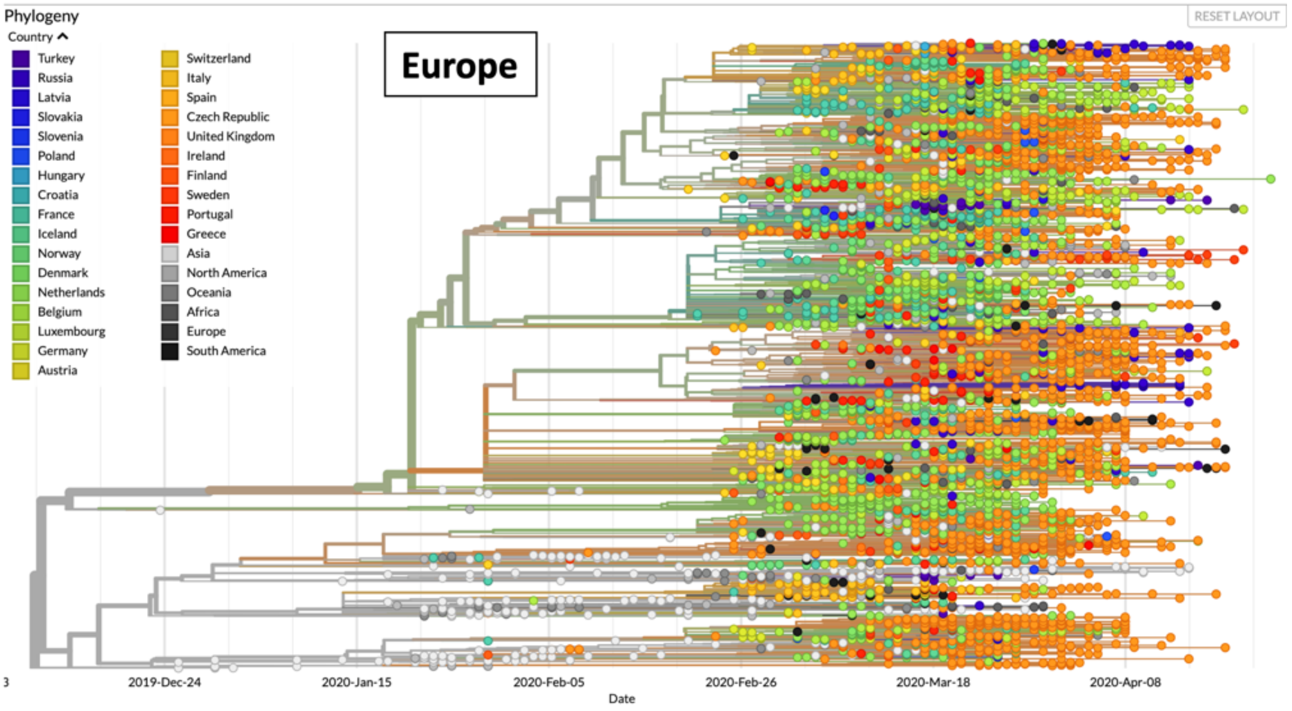

Accumulation of genomic mutations of SARS-CoV-2 over time in Europe. Each data-point is represented as a bead, whereby each bead corresponds to a specific set of virus mutations (mutation haplotype). Beads and bead sequences are color-coded, according to the country where the virus sample was isolated from (upper left). The 'beads-on-a-string' plots link successions of viral mutations, i.e. mutation haplotypes that acquired additional mutations over time. Phylogeny trees for such mutations are presented, that draw distinct evolutionary branches of SARS-CoV-2 over calendar dates (bottom) (nextstrain.org/ncov/europe?branchLabel=aa).

**Figure S4. Genomic mutations of SARS-CoV-2 - Spain.**

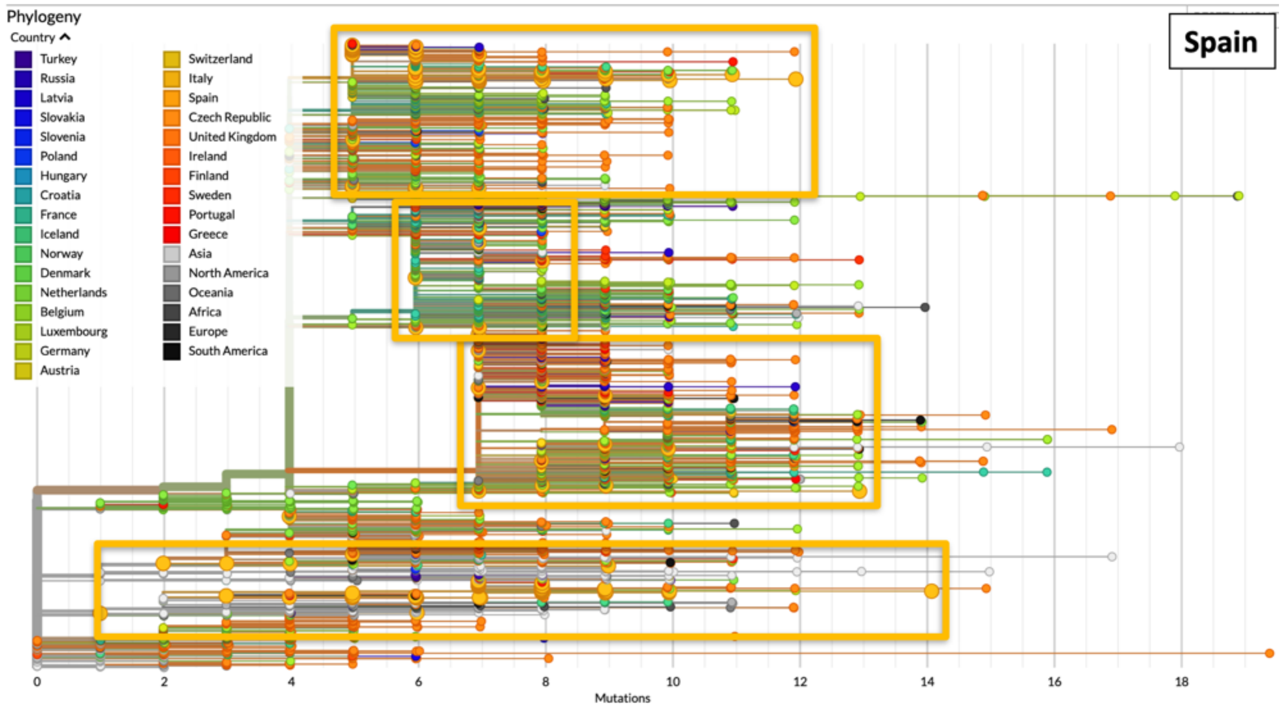

Accumulation of genomic mutations of SARS-CoV-2 over time in Spain is shown as strings of dark-yellow large dots. Each data-point is represented as a bead, whereby each bead corresponds to a specific set of virus mutations (mutation haplotype). The 'beads-on-a-string' plots link successions of viral mutations, i.e. mutation haplotypes that acquired additional mutations over time. Phylogeny trees for such mutations are presented, that draw distinct evolutionary branches of SARS-CoV-2 (nextstrain.org/ncov/europe?branchLabel=aa). Dark-yellow rectangles enclose successions of viral mutations in Spain in each phylogenetic branch.

The number of accumulated mutations in individual viral isolates is indicated at the bottom.

Color codes by country are shown in the upper left.

**Figure S5. Genomic mutations of SARS-CoV-2 - Italy.**

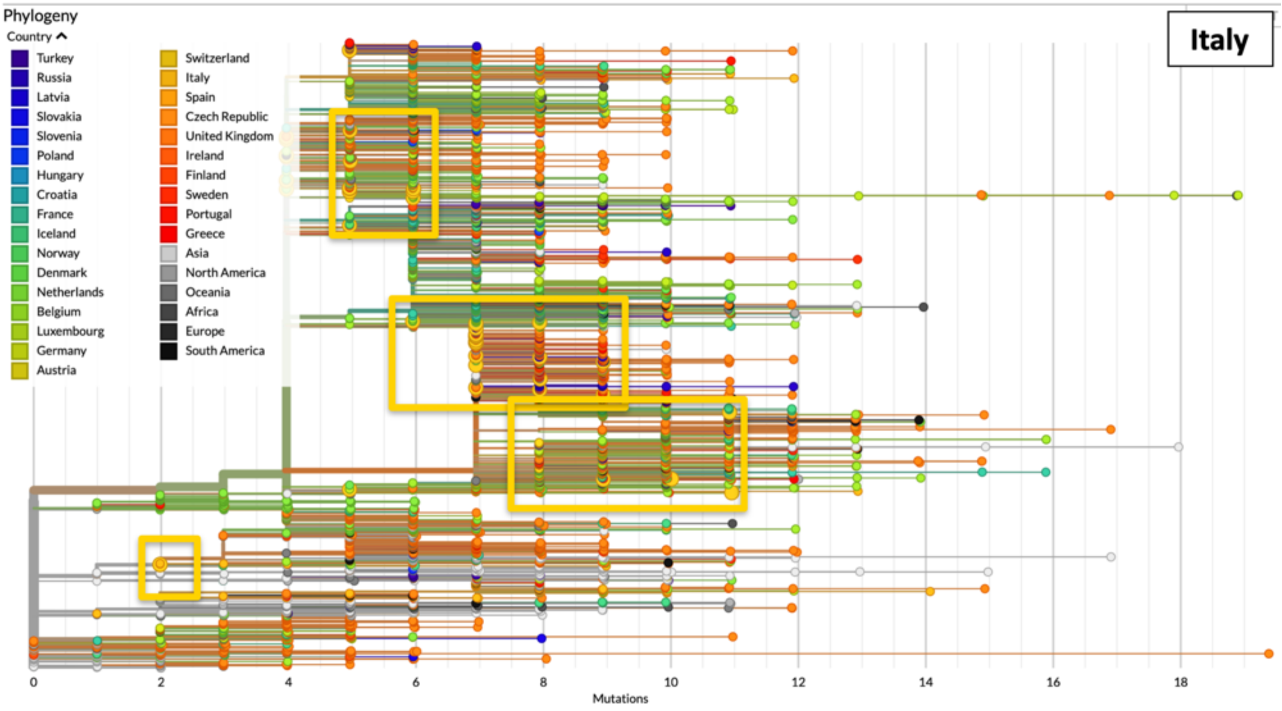

Accumulation of genomic mutations of SARS-CoV-2 over time in Italy is shown as strings of yellow large dots. Each data-point is represented as a bead, whereby each bead corresponds to a specific set of virus mutations (mutation haplotype). The 'beads-on-a-string' plots link successions of viral mutations, i.e. mutation haplotypes that acquired additional mutations over time. Phylogeny trees for such mutations are presented, that draw distinct evolutionary branches of SARS-CoV-2 (nextstrain.org/ncov/europe?branchLabel=aa). Yellow rectangles enclose successions of viral mutations in Italy in each phylogenetic branch.

The number of accumulated mutations in individual viral isolates is indicated at the bottom.

Color codes by country are shown in the upper left.

**Figure S6. Genomic mutations of SARS-CoV-2 - Sweden.**

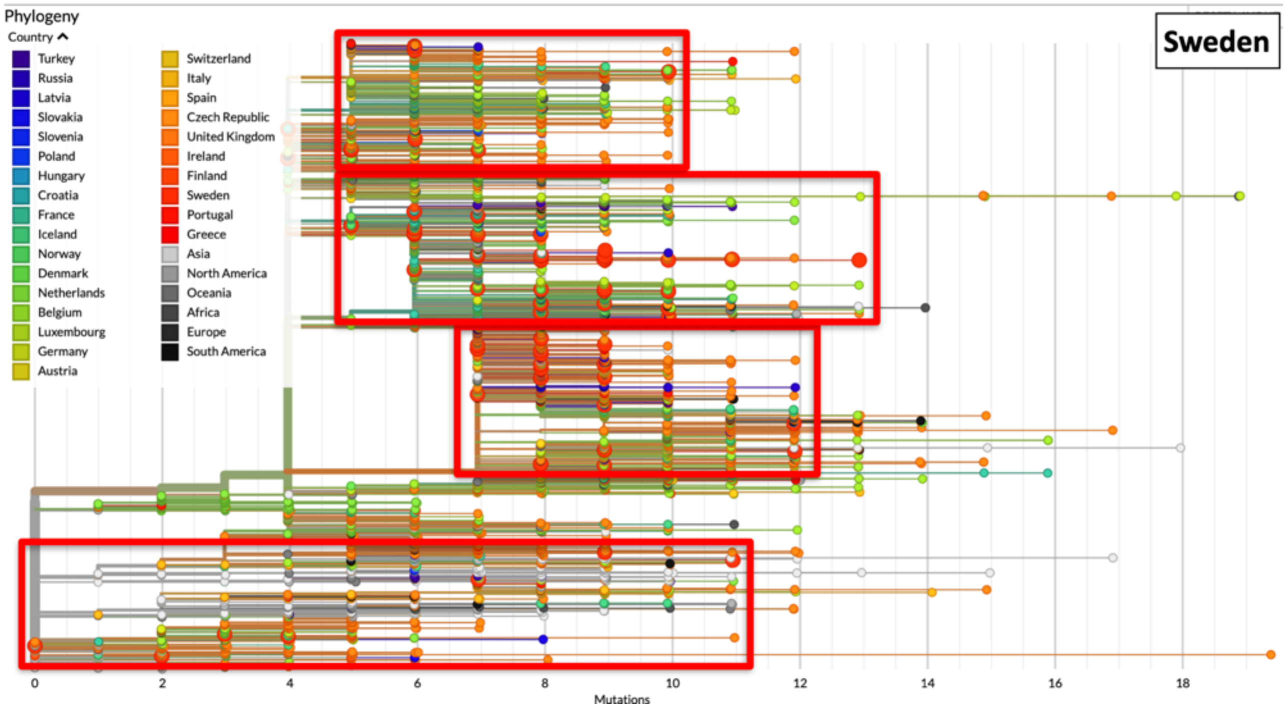

Accumulation of genomic mutations of SARS-CoV-2 over time in Sweden is shown as strings of red large dots. Each data-point is represented as a bead, whereby each bead corresponds to a specific set of virus mutations (mutation haplotype). The 'beads-on-a-string' plots link successions of viral mutations, i.e. mutation haplotypes that acquired additional mutations over time. Phylogeny trees for such mutations are presented, that draw distinct evolutionary branches of SARS-CoV-2 (nextstrain.org/ncov/europe?branchLabel=aa). Red rectangles enclose successions of viral mutations in Sweden in each phylogenetic branch.

The number of accumulated mutations in individual viral isolates is indicated at the bottom.

Color codes by country are shown in the upper left.

**Figure S7. Genomic mutations of SARS-CoV-2 - The Netherlands.**

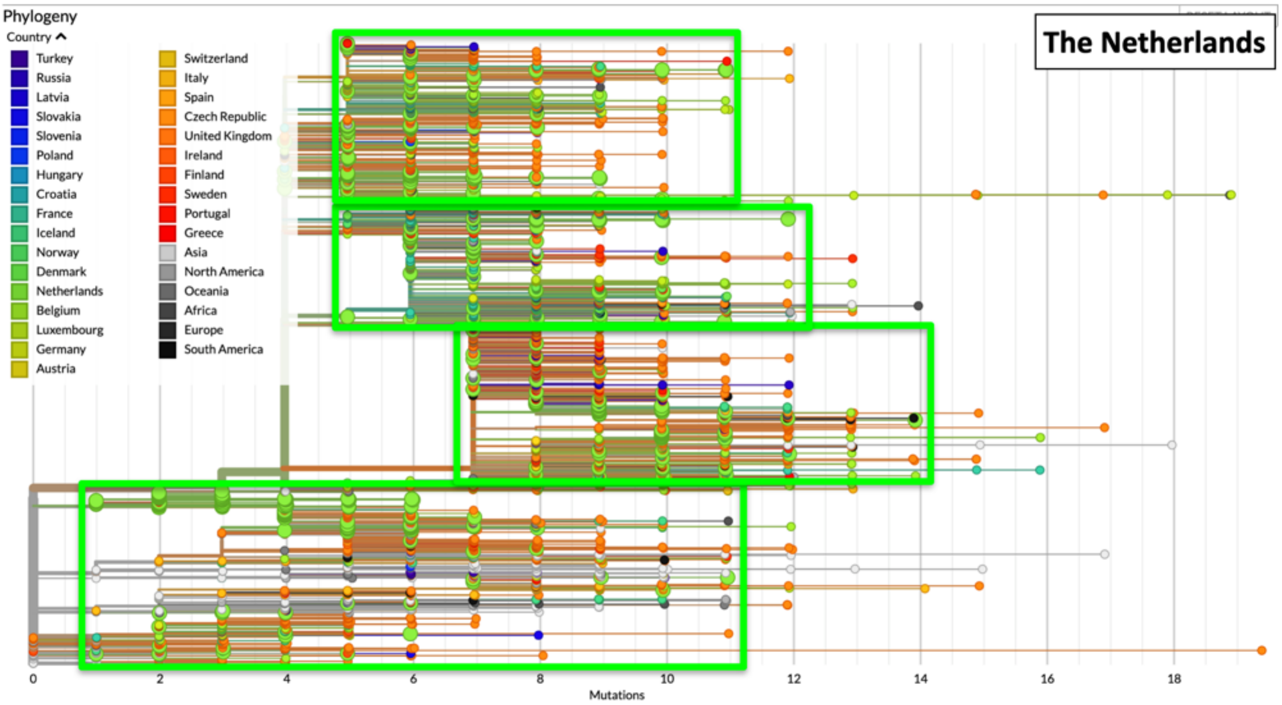

Accumulation of genomic mutations of SARS-CoV-2 over time in The Netherlands is shown as strings of bright-green large dots. Each data-point is represented as a bead, whereby each bead corresponds to a specific set of virus mutations (mutation haplotype). The 'beads-on-a-string' plots link successions of viral mutations, i.e. mutation haplotypes that acquired additional mutations over time. Phylogeny trees for such mutations are presented, that draw distinct evolutionary branches of SARS-CoV-2 (nextstrain.org/ncov/europe?branchLabel=aa). Bright-green rectangles enclose successions of viral mutations in The Netherlands in each phylogenetic branch.

The number of accumulated mutations in individual viral isolates is indicated at the bottom.

Color codes by country are shown in the upper left.

**Figure S8. Genomic mutations of SARS-CoV-2 - Belgium.**

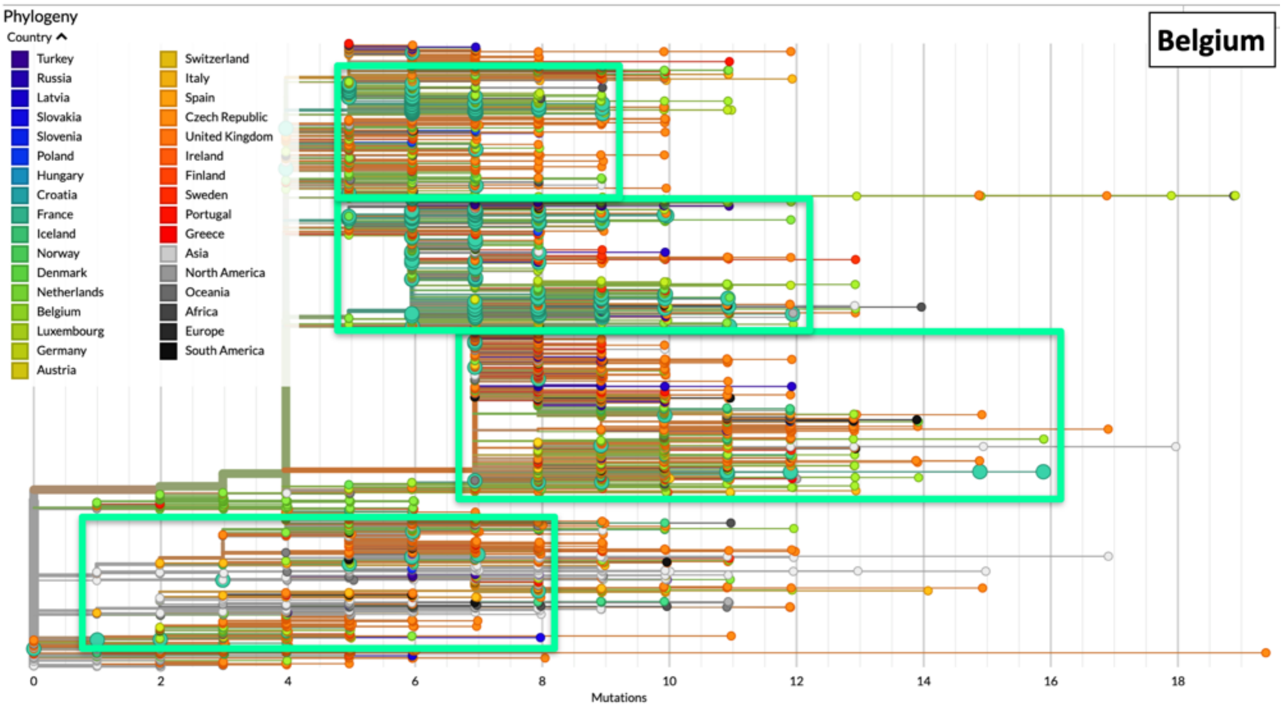

Accumulation of genomic mutations of SARS-CoV-2 over time in Belgium is shown as strings of light-green large dots. Each data-point is represented as a bead, whereby each bead corresponds to a specific set of virus mutations (mutation haplotype). The 'beads-on-a-string' plots link successions of viral mutations, i.e. mutation haplotypes that acquired additional mutations over time. Phylogeny trees for such mutations are presented, that draw distinct evolutionary branches of SARS-CoV-2 (nextstrain.org/ncov/europe?branchLabel=aa). Light-green rectangles enclose successions of viral mutations in Belgium in each phylogenetic branch.

The number of accumulated mutations in individual viral isolates is indicated at the bottom.

Color codes by country are shown in the upper left.

**Figure S9. Genomic mutations of SARS-CoV-2 - France.**

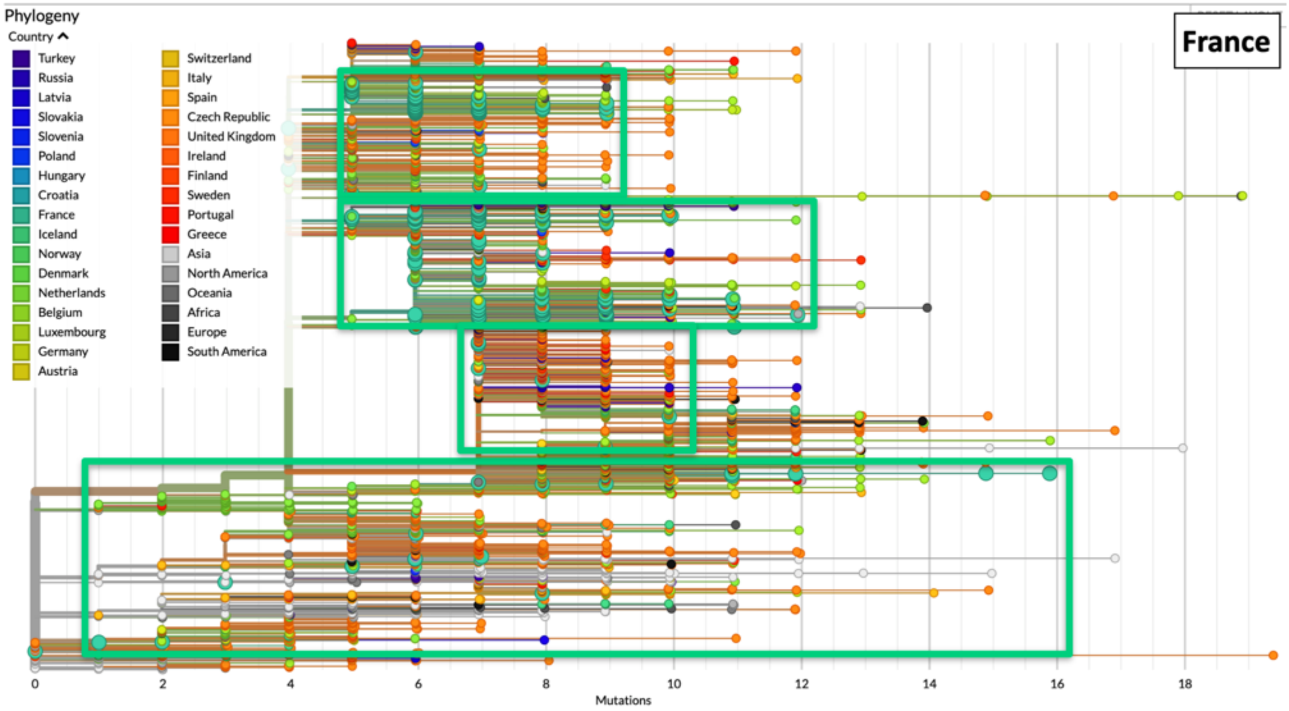

Accumulation of genomic mutations of SARS-CoV-2 over time in France is shown as strings of deep-green large dots. Each data-point is represented as a bead, whereby each bead corresponds to a specific set of virus mutations (mutation haplotype). The 'beads-on-a-string' plots link successions of viral mutations, i.e. mutation haplotypes that acquired additional mutations over time. Phylogeny trees for such mutations are presented, that draw distinct evolutionary branches of SARS-CoV-2 (nextstrain.org/ncov/europe?branchLabel=aa). Deep-green rectangles enclose successions of viral mutations in France in each phylogenetic branch.

The number of accumulated mutations in individual viral isolates is indicated at the bottom.

Color codes by country are shown in the upper left.

**Figure S10. Progression of COVID-19 over Italy -** Northern Provinces ***.**

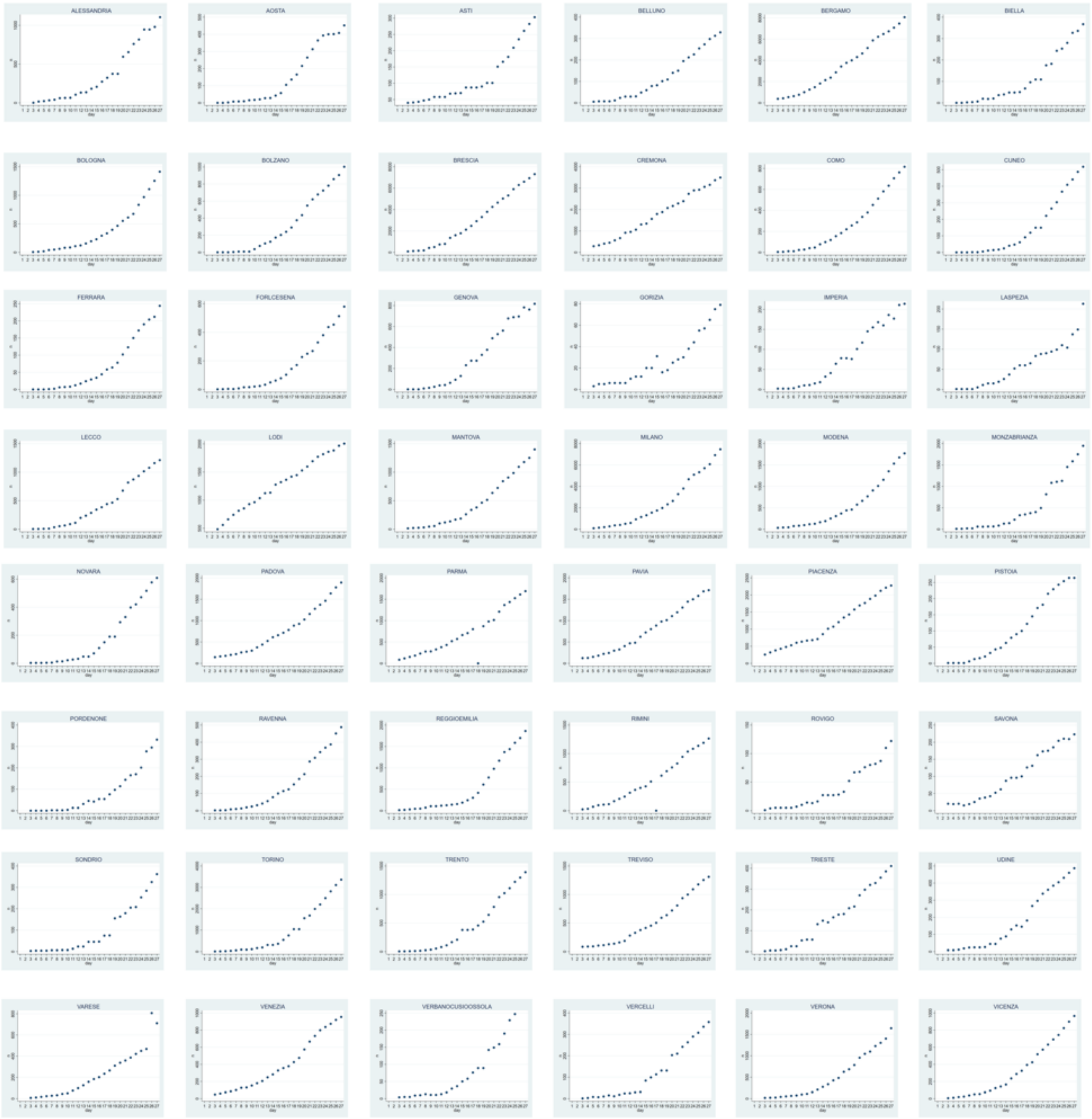

*: Case incidence in the indicated municipalities is plotted versus time (days, from March 3 to March 27, 2020).
For comparison purposes, all graphs were normalized versus the highest number of cases per province. COVID-19 doubling times were computed on non-normalized, absolute numbers of infection cases.
Graphs are in alphabetical order by province name.

**Figure S11. Progression of COVID-19 over Italy -** Central & Southern Provinces***.**

Central

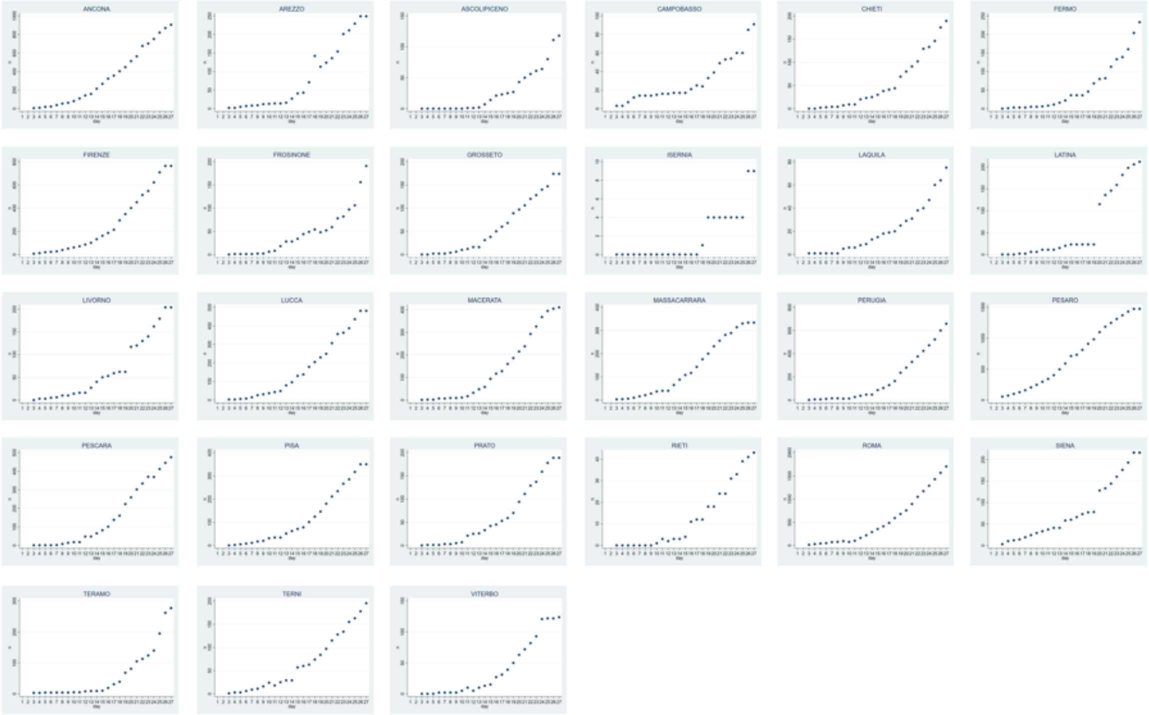

Southern

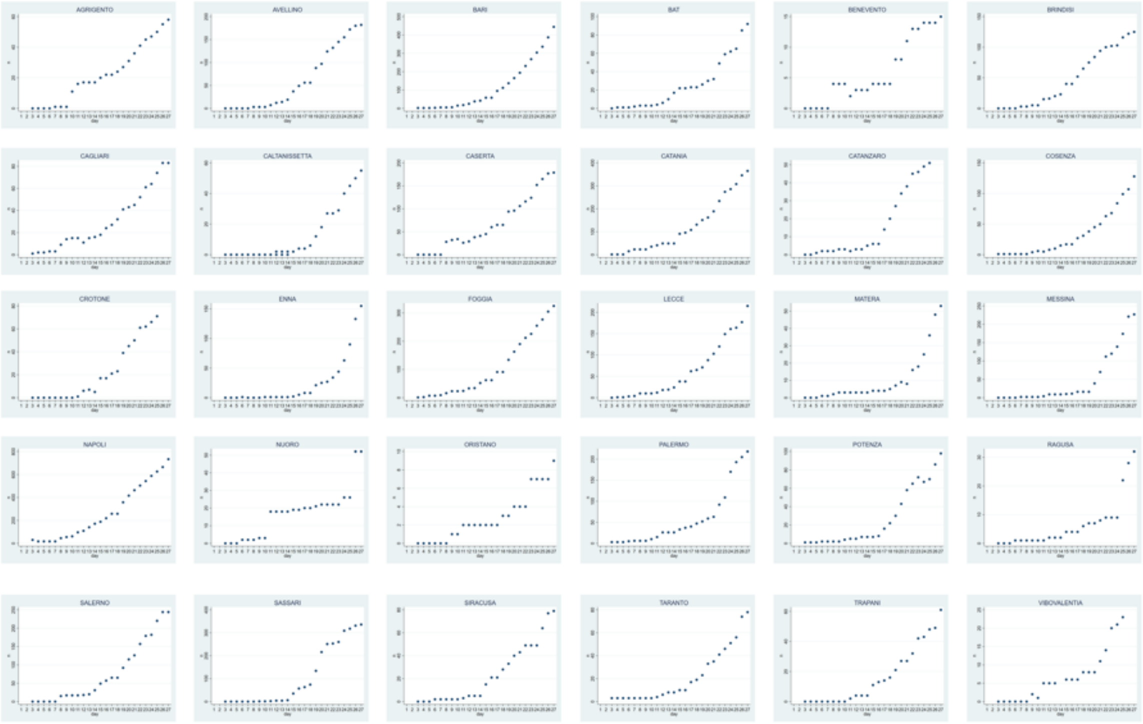

*: Case incidence in the indicated municipalities is plotted versus time (days, from March 3 to March 27, 2020). For comparison purposes, all graphs were normalized versus the highest number of cases per Province. COVID-19 doubling times were computed on non-normalized, absolute numbers. Graphs are in alphabetical order by province name.

**Figure S12. Progression of COVID-19 over Italy – global data by region*.**

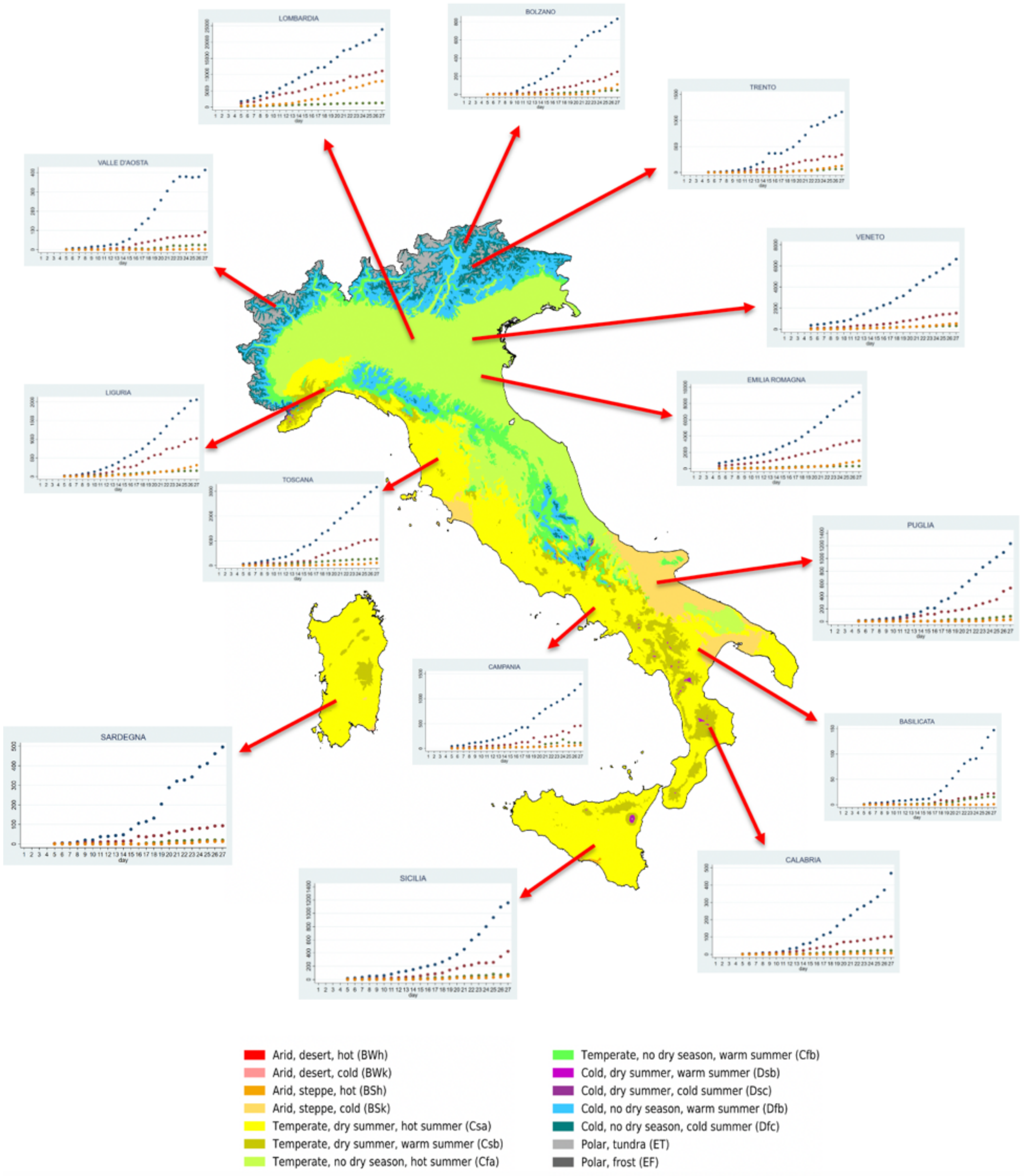

Köppen–Geiger climate classification map. Compounded data of COVID-19 spreading in individual Italy's regions versus time (days, from March 3 to March 27, 2020) are overlaid over the country's climate areas. (*insets*) Dark gray dots: SARS-CoV-2-positive cases; brown dots: hospitalized cases; green dots: intensive-care unit cases; orange dots: recovered cases.
(*bottom*) color codes for climate areas classification.

**Figure S13. Progression of COVID-19 over Spain*.**

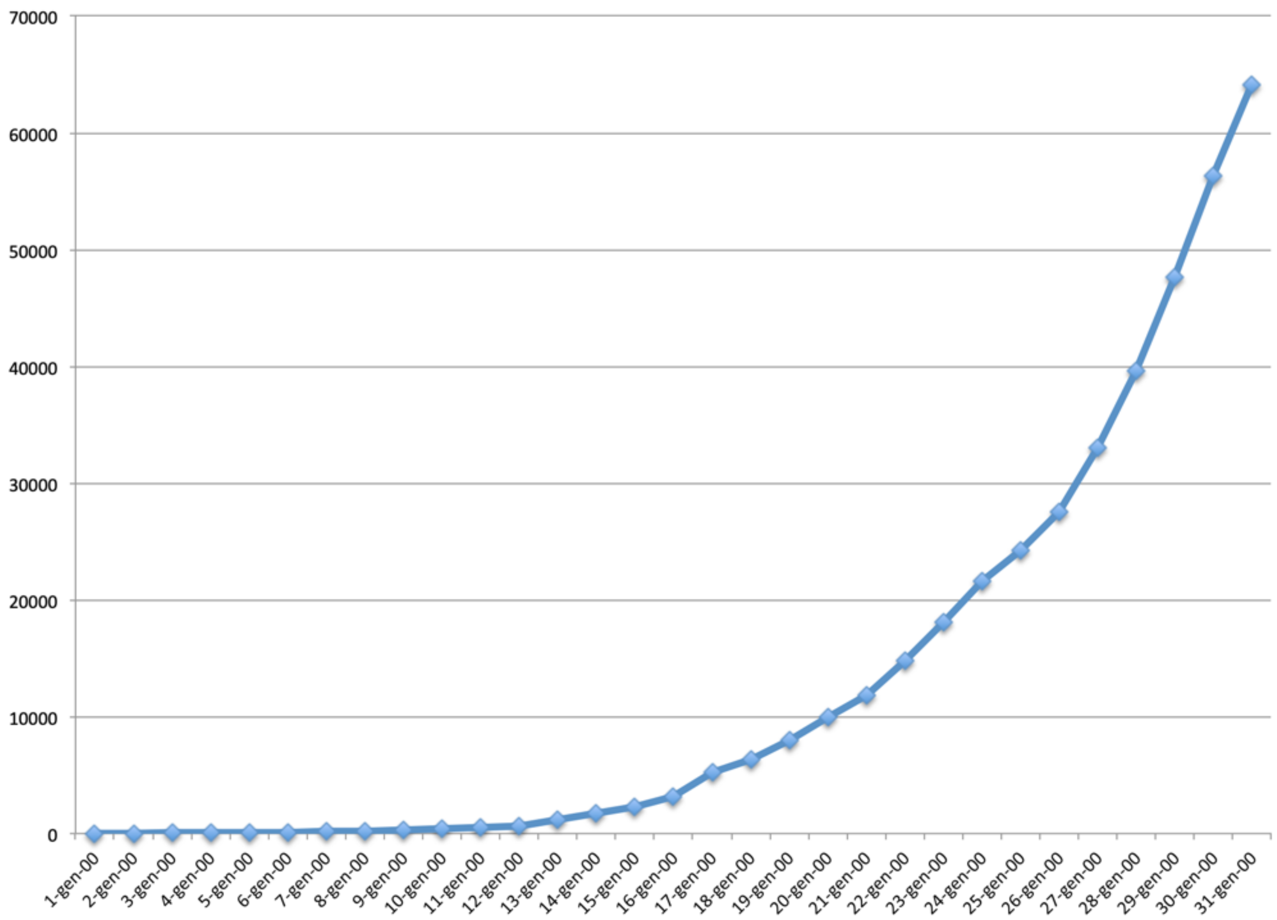

*: cumulative case incidence in Spain is plotted versus calendar dates (March 1-31, 2020).

**Figure S14. Progression of COVID-19 over Norway*.**

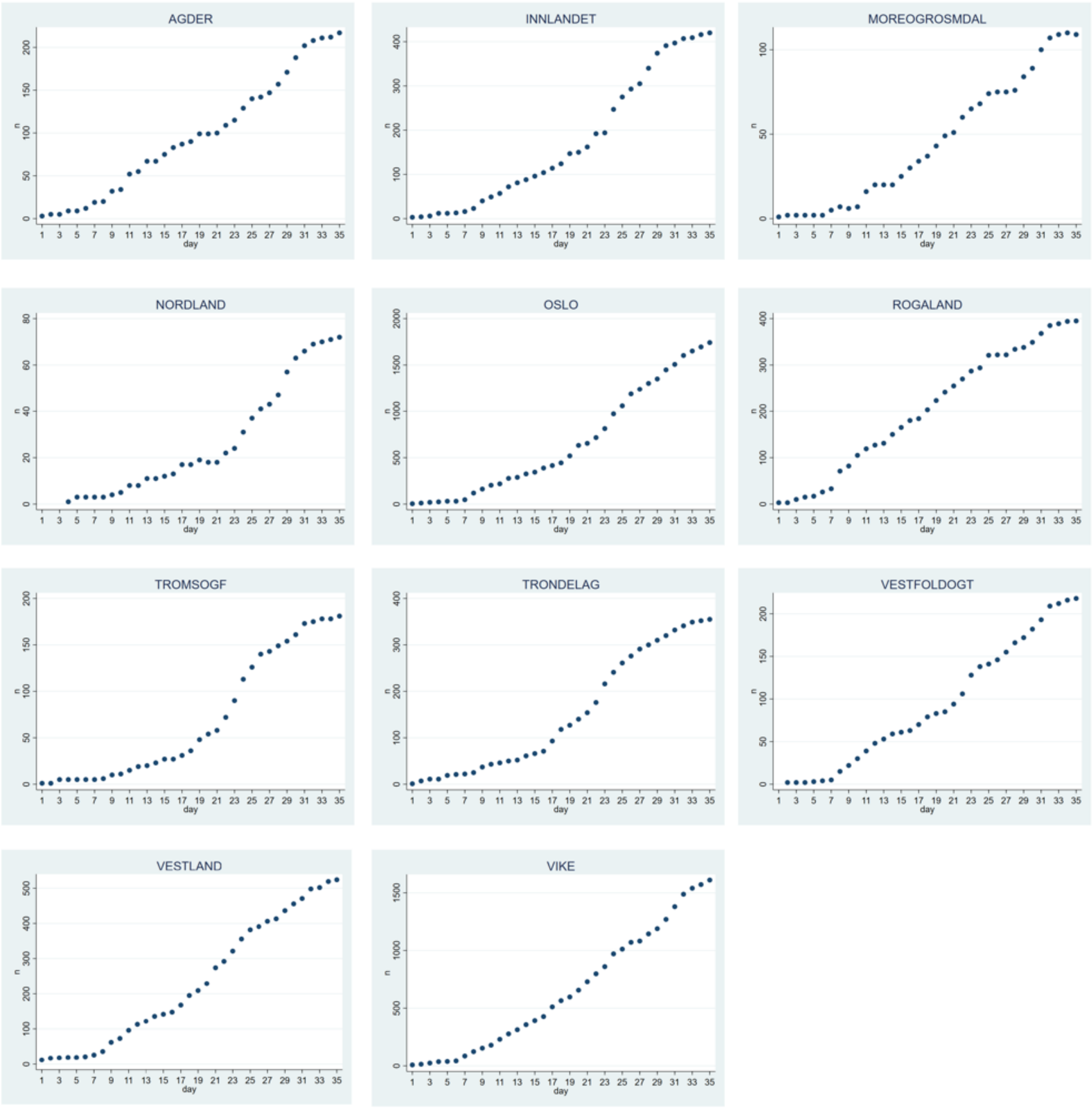

*: cumulative case incidence in the indicated municipalities is plotted versus time (days, from March 1 to April 7, 2020).

**Figure S15. Progression of COVID-19 over Finland*.**

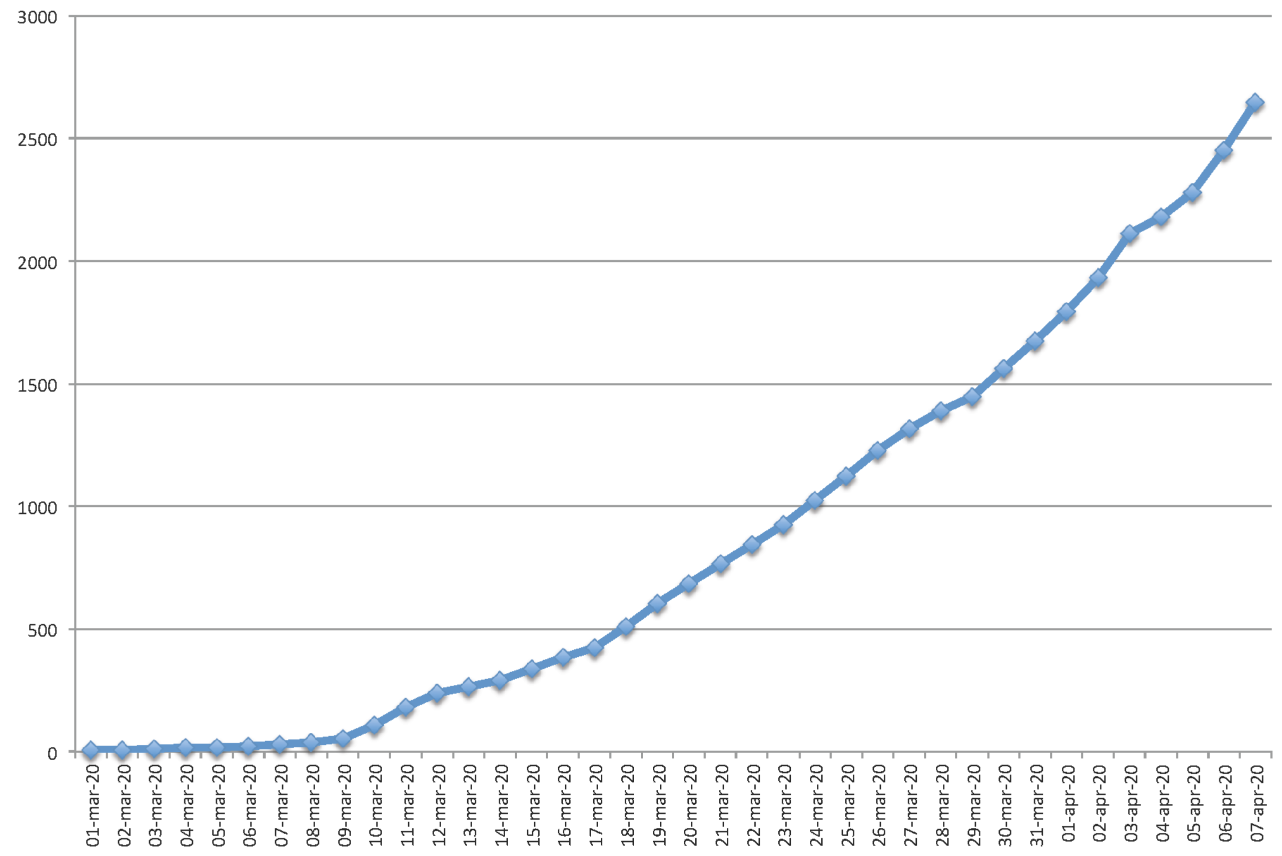

*: cumulative case incidence in the indicated municipalities is plotted versus calendar dates (March 1-April 7, 2020).

**Figure S16. Progression of COVID-19 over Sweden*.**

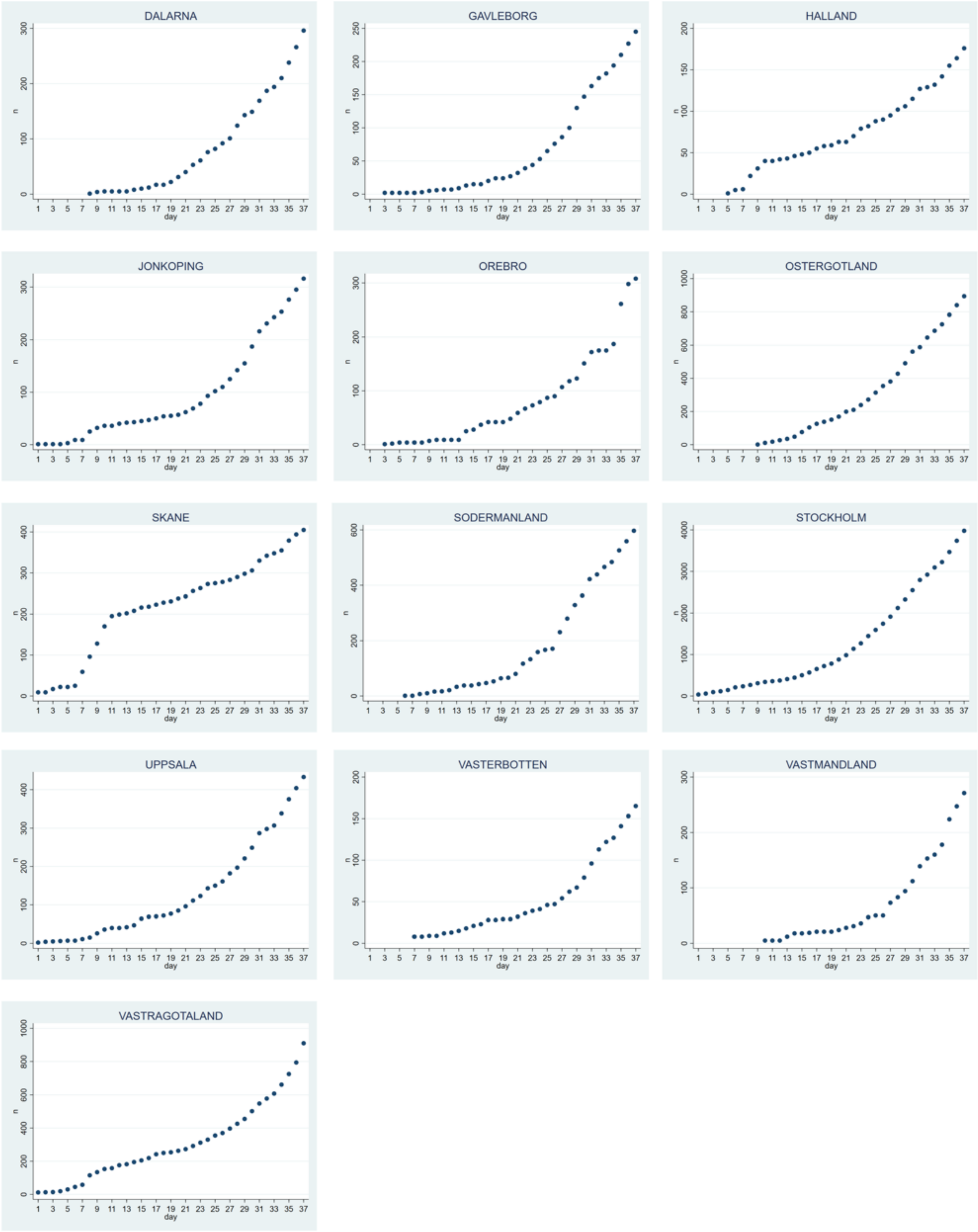

*: cumulative case incidence in the indicated municipalities is plotted versus time (days, March 1-March 23, 2020).

**Figure S17. Progression of COVID-19 over France, Germany and UK*.**

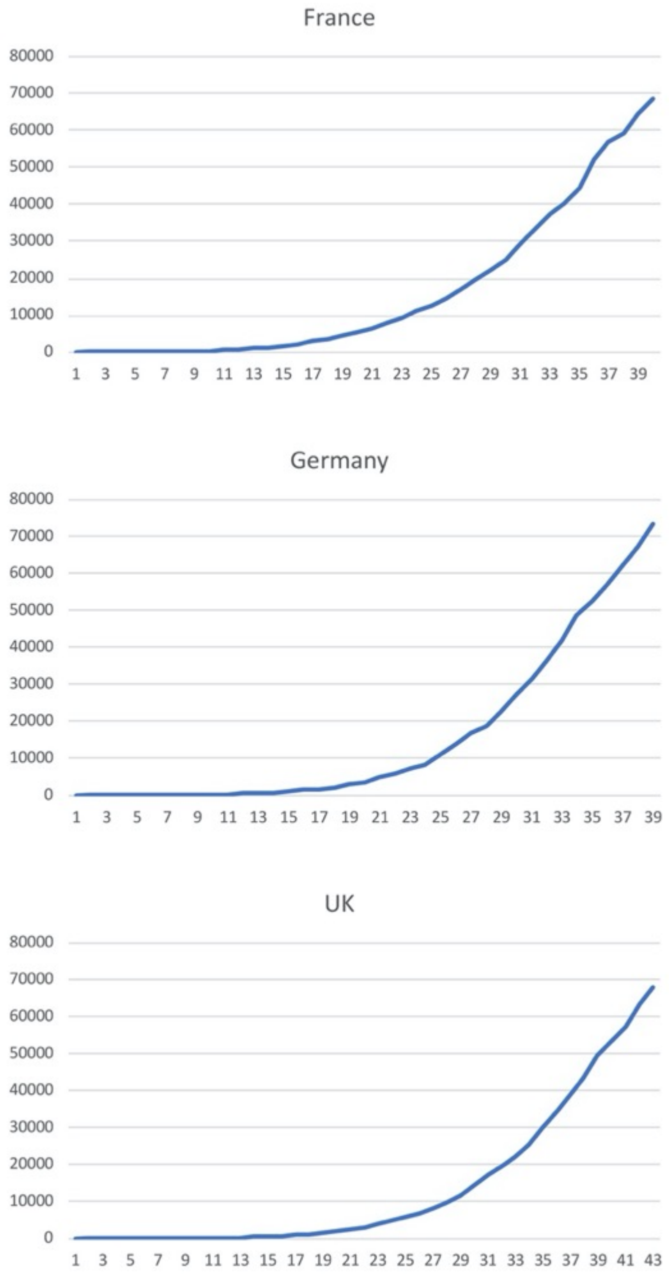

*: Laboratory-confirmed infection cases in Europe cases were retrieved by country before landmark dates: France (dashboard.covid19.data.gouv.fr/vue-d-ensemble?locatio n=FRA; April 4th 2020); UK (www.nhs.uk/; April 9th 2020); Germany (corona.rki.de; April 2nd 2020). Cumulative case incidence is plotted over time (days).

**Supplemental online Table 1**. COVID-19 spreading in Central and South America.

| | **Argentina** | **Brazil** | **Chile** | **Colombia** | **Costa Rica** | **Dominican Republic** | **Ecuador** | **Mexico** | **Panama** | **Peru** | **Uruguay** | **Venezuela** | **Trinidad and Tobago** |
|---|---|---|---|---|---|---|---|---|---|---|---|---|---|
| March 1* | | 2 | | | | 1 | 1 | 2 | | | | | |
| March 2 | | 2 | | | | 1 | 1 | 5 | | | | | |
| March 3 | | 2 | 1 | | | 1 | 6 | 5 | | | | | |
| March 4 | 1 | 2 | 1 | | | 1 | 7 | 5 | | | | | |
| March 5 | 1 | 3 | 1 | | 1 | 1 | 7 | 5 | | | | | |
| March 6 | 1 | 7 | 1 | 1 | 1 | 1 | 16 | 5 | | | | | |
| March 7 | 2 | 13 | 5 | 1 | 1 | 1 | 14 | 5 | | 1 | | | |
| March 8 | 9 | 19 | 5 | 1 | 5 | 1 | 14 | 7 | | 6 | | | |
| March 9 | 12 | 25 | 10 | 1 | 9 | 1 | 15 | 7 | | 6 | | | |
| March 10 | 12 | 25 | 13 | 3 | 9 | 5 | 15 | 7 | 1 | 9 | | | |
| March 11 | 17 | 34 | 17 | 3 | 13 | 5 | 15 | 7 | 8 | 11 | | | |
| March 12 | 19 | 52 | 23 | 9 | 13 | 5 | 17 | 11 | 10 | 17 | | | |
| March 13 | 31 | 77 | 33 | 9 | 22 | 5 | 17 | 12 | 14 | 22 | | | |
| March 14 | 34 | 98 | 43 | 16 | 23 | 5 | 23 | 26 | 27 | 28 | | 2 | 1 |
| March 15 | 45 | 121 | 61 | 24 | 23 | 5 | 23 | 41 | 27 | 43 | | 2 | 1 |
| March 16 | 56 | 200 | 75 | 24 | 23 | 5 | 37 | 53 | 43 | 71 | 4 | 2 | 2 |
| March 17 | 65 | 234 | 156 | 45 | 41 | 21 | 58 | 53 | 69 | 86 | 6 | 33 | 5 |
| March 18 | 65 | 234 | 156 | 45 | 41 | 21 | 58 | 82 | 69 | 86 | 6 | 33 | 5 |
| March 19 | 79 | 291 | 238 | 93 | 50 | 21 | 155 | 93 | 86 | 145 | 29 | 36 | 7 |
| March 20 | 97 | 428 | 342 | 108 | 87 | 34 | 199 | 118 | 109 | 234 | 79 | 36 | 9 |
| March 21 | 128 | 621 | 434 | 145 | 113 | 72 | 367 | 164 | 137 | 234 | 94 | 36 | 9 |
| March 22 | 158 | 904 | 434 | 196 | 113 | 72 | 506 | 164 | 137 | 318 | 94 | 36 | 9 |
| March 23 | 225 | 904 | 632 | 196 | 117 | 72 | 532 | 251 | 245 | 318 | 135 | 70 | 50 |

*: cases were recorded from March 1 to March 23, 2020.

**Supplemental online Table 2**: COVID-19 in Middle-East, Africa, and Gulf countries.

| | Iran | Pakistan | Saudi Arabia | Qatar | Bahrain | Egypt | Lebanon | Iraq | Kuwait | UAE | Morocco | Jordan | Tunisia | Oman | Afghanistan | South Africa | Algeria | Burkina Faso | Senegal | DRC | Cameroon |
|---|---|---|---|---|---|---|---|---|---|---|---|---|---|---|---|---|---|---|---|---|---|
| March 1* | 593 | 4 | | 1 | 40 | 1 | 2 | 13 | 45 | 19 | | | | 6 | 1 | | | | | | |
| March 2 | 978 | 4 | | 3 | 47 | 2 | 10 | 19 | 56 | 21 | | | | 6 | 1 | | | | | | |
| March 3 | 1501 | 5 | 1 | 7 | 49 | 2 | 13 | 26 | 56 | 21 | 1 | 1 | 1 | 6 | 1 | | 5 | | | | |
| March 4 | 2336 | 5 | 1 | 8 | 49 | 2 | 13 | 31 | 56 | 27 | 1 | 1 | 1 | 12 | 1 | | 5 | | | | |
| March 5 | 2922 | 5 | 2 | 8 | 49 | 2 | 13 | 36 | 58 | 27 | 2 | 1 | 1 | 15 | 1 | | 12 | | | | |
| March 6 | 3513 | 5 | 8 | 8 | 49 | 3 | 16 | 36 | 58 | 27 | 2 | 1 | 1 | 16 | 1 | | 12 | | 4 | | 1 |
| March 7 | 4747 | 5 | 8 | 11 | 49 | 3 | 22 | 44 | 58 | 45 | 2 | 1 | 1 | 16 | 1 | 1 | 17 | | 4 | | 2 |
| March 8 | 5823 | 5 | 7 | 12 | 56 | 48 | 28 | 54 | 62 | 45 | 2 | 1 | 1 | 16 | 4 | 2 | 17 | | 4 | | 2 |
| March 9 | 6566 | 6 | 15 | 15 | 79 | 55 | 32 | 60 | 64 | 45 | 2 | 1 | 2 | 16 | 4 | 3 | 20 | | 4 | | 2 |
| March 10 | 7161 | 16 | 15 | 18 | 109 | 59 | 41 | 61 | 65 | 59 | 2 | 1 | 2 | 18 | 4 | 7 | 20 | | 4 | | 2 |
| March 11 | 8042 | 16 | 20 | 24 | 110 | 59 | 41 | 61 | 69 | 74 | 3 | 1 | 6 | 18 | 4 | 7 | 20 | 2 | 4 | 1 | 2 |
| March 12 | 9000 | 19 | 21 | 262 | 189 | 67 | 66 | 70 | 80 | 74 | 5 | 1 | 6 | 18 | 7 | 13 | 25 | 2 | 4 | 1 | 2 |
| March 13 | 10075 | 20 | 21 | 262 | 195 | 67 | 66 | 70 | 80 | 85 | 6 | 1 | 7 | 18 | 7 | 17 | 25 | 2 | 10 | 1 | 2 |
| March 14 | 11364 | 21 | 62 | 262 | 210 | 93 | 77 | 93 | 100 | 85 | 7 | 1 | 16 | 19 | 7 | 17 | 26 | 2 | 10 | 2 | 2 |
| March 15 | 12729 | 28 | 103 | 337 | 211 | 93 | 93 | 93 | 112 | 85 | 18 | 1 | 16 | 20 | 10 | 38 | 37 | 3 | 21 | 2 | 3 |
| March 16 | 14991 | 52 | 103 | 401 | 221 | 126 | 99 | 124 | 112 | 98 | 28 | 6 | 18 | 22 | 16 | 51 | 49 | 3 | 26 | 2 | 3 |
| March 17 | 14991 | 187 | 133 | 439 | 229 | 166 | 109 | 124 | 130 | 98 | 38 | 35 | 24 | 21 | 21 | 62 | 60 | 15 | 27 | 3 | 5 |
| March 18 | 16169 | 187 | 171 | 442 | 237 | 166 | 120 | 154 | 130 | 98 | 38 | 35 | 24 | 24 | 22 | 62 | 60 | 20 | 27 | 3 | 5 |
| March 19 | 17361 | 241 | 238 | 442 | 258 | 196 | 133 | 164 | 142 | 113 | 49 | 52 | 29 | 33 | 22 | 116 | 72 | 26 | 36 | 7 | 10 |
| March 20 | 18407 | 302 | 238 | 452 | 269 | 210 | 149 | 177 | 148 | 140 | 61 | 56 | 39 | 39 | 22 | 150 | 82 | 40 | 38 | 14 | 15 |
| March 21 | 19644 | 461 | 274 | 460 | 285 | 256 | 163 | 193 | 159 | 140 | 74 | 69 | 54 | 48 | 24 | 205 | 94 | 40 | 38 | 14 | 22 |
| March 22 | 20610 | 495 | 392 | 470 | 306 | 285 | 206 | 214 | 176 | 153 | 86 | 84 | 60 | 52 | 24 | 240 | 94 | 72 | 56 | 23 | 27 |
| March 23 | 21638 | 748 | 511 | 494 | 337 | 327 | 248 | 233 | 189 | 153 | 115 | 112 | 75 | 55 | 40 | 274 | 201 | 75 | 67 | 30 | 40 |

*: cases were recorded from March 1 to March 23, 2020. UAE: United Arab Emirates. DRC: Democratic Republic of the Congo.

**Supplemental online Table 3**. COVID-19 doubling time by province *.

| | Doubling | Geographical |
|---|---|---|
| AGRIGENTO | 6,67 | South |
| ALESSANDRIA | 5,55 | North |
| ANCONA | 6,84 | Center |
| AOSTA | 6,29 | North |
| AREZZO | 5,86 | Center |
| ASCOLIPICENO | 5,19 | Center |
| ASTI | 5,41 | North |
| AVELLINO | 6,06 | South |
| BARI | 4,90 | South |
| BAT | 3,97 | South |
| BELLUNO | 6,10 | North |
| BENEVENTO | 6,25 | South |
| BERGAMO | 9,65 | North |
| BIELLA | 5,17 | North |
| BOLOGNA | 4,97 | North |
| BOLZANO | 6,12 | North |
| BRESCIA | 8,00 | North |
| BRINDISI | 7,54 | South |
| CAGLIARI | 6,55 | South |
| CALTANISSETTA | 4,50 | South |
| CAMPOBASSO | 6,33 | Center |
| CASERTA | 6,40 | South |
| CATANIA | 5,73 | South |
| CATANZARO | 6,21 | South |
| CHIETI | 5,64 | Center |
| COMO | 5,64 | North |
| COSENZA | 4,10 | South |
| CREMONA | 10,94 | North |
| CROTONE | 6,22 | South |
| CUNEO | 5,01 | North |
| ENNA | 1,95 | South |
| FERMO | 5,00 | Center |
| FERRARA | 5,00 | North |
| FIRENZE | 5,89 | Center |
| FOGGIA | 5,84 | South |
| FORLCESENA | 6,00 | North |
| FROSINONE | 4,86 | Center |
| GENOVA | 6,88 | North |
| GORIZIA | 4,69 | North |
| GROSSETO | 6,74 | Center |
| IMPERIA | 7,50 | North |
| ISERNIA | 6,67 | Center |
| LAQUILA | 4,50 | Center |
| LASPEZIA | 7,81 | North |
| LATINA | 5,17 | Center |
| LECCE | 5,35 | South |
| LECCO | 5,95 | North |
| LIVORNO | 5,50 | Center |
| LODI | 15,60 | North |
| LUCCA | 6,48 | Center |
| MACERATA | 5,60 | Center |
| MANTOVA | 6,39 | North |
| MASSACARRARA | 7,33 | Center |
| MATERA | 2,00 | South |
| MESSINA | 3,60 | South |
| MILANO | 6,38 | North |
| MODENA | 5,00 | North |
| MONZABRIANZA | 5,07 | North |
| NAPOLI | 6,45 | South |

| | | |
|---|---|---|
| NOVARA | 5,33 | North |
| NUORO | 14,33 | South |
| ORISTANO | 5,50 | South |
| PADOVA | 7,63 | North |
| PALERMO | 2,74 | South |
| PARMA | 6,12 | North |
| PAVIA | 9,15 | North |
| PERUGIA | 5,42 | Center |
| PESARO | 9,81 | Center |
| PESCARA | 6,27 | Center |
| PIACENZA | 9,20 | North |
| PISA | 5,63 | Center |
| PISTOIA | 6,78 | Center |
| PORDENONE | 4,18 | North |
| POTENZA | 5,62 | South |
| PRATO | 5,21 | Center |
| RAGUSA | 0,85 | South |
| RAVENNA | 5,70 | North |
| REGGIOEMILIA | 4,90 | North |
| RIETI | 4,75 | Center |
| RIMINI | 7,07 | North |
| ROMA | 5,53 | Center |
| ROVIGO | 6,45 | North |
| SALERNO | 5,22 | South |
| SASSARI | 5,70 | South |
| SAVONA | 7,81 | North |
| SIENA | 5,63 | Center |
| SIRACUSA | 6,20 | South |
| SONDRIO | 6,16 | North |
| TARANTO | 5,50 | South |
| TERAMO | 4,31 | Center |
| TERNI | 6,25 | Center |
| TORINO | 5,29 | North |
| TRAPANI | 5,50 | South |
| TRENTO | 5,26 | North |
| TREVISO | 7,03 | North |
| TRIESTE | 7,83 | North |
| UDINE | 6,59 | North |
| VARESE | 8,00 | North |
| VENEZIA | 6,78 | North |
| VERBANOCUSIOOSSOLA | 5,35 | North |
| VERCELLI | 5,68 | North |
| VERONA | 6,57 | North |
| VIBOVALENTIA | 3,83 | South |
| VICENZA | 6,39 | North |
| VITERBO | 5,15 | Center |
| *: The COVID-19 doubling time was computed on province-base | | |

**Supplemental online Table 4**. COVID-19 spreading in Scandinavia by county *.

| Sweden | | | | Norway | |
|---|---|---|---|---|---|
| | Doubling time | | | | Doubling time |
| DALARNA | 7,17 | | | AGDER | 13,06 |
| GAVLEBORG | 8,25 | | | INNLANDET | 11,70 |
| HALLAND | 12,00 | | | MOREOGROSMDAL | 13,61 |
| JONKOPING | 7,91 | | | NORDLAND | 10,17 |
| OREBRO | 6,86 | | | OSLO | 11,64 |
| OSTERGOTLAND | 8,70 | | | ROGALAND | 17,29 |
| SKANE | 23,92 | | | TROMSOGF | 11,98 |
| SODERMANLAND | 8,62 | | | TRONDELAG | 12,96 |
| STOCKHOLM | 9,64 | | | VESTFOLDOGT | 12,86 |
| UPPSALA | 8,19 | | | VESTLAND | 14,27 |
| VASTERBOTTEN | 6,79 | | | VIKE | 12,88 |
| VASTMANDLAND | 6,13 | | | | |
| VASTRAGOTALAND | 8,00 | | | | |

*: Landmark dates were the 9th of April for Sweden and the 7th of April for Norway.

**Supplemental online Table 5**. COVID-19 cumulative incidence by date in France.

| DATE | TOTAL CASES |
|---|---|
| 25/02/20 | 13 |
| 26/02/20 | 18 |
| 27/02/20 | 38 |
| 28/02/20 | 57 |
| 29/02/20 | 100 |
| 01/03/20 | 130 |
| 02/03/20 | 191 |
| 03/03/20 | 212 |
| 04/03/20 | 285 |
| 05/03/20 | 423 |
| 06/03/20 | 613 |
| 07/03/20 | 949 |
| 08/03/20 | 1126 |
| 09/03/20 | 1412 |
| 10/03/20 | 1784 |
| 11/03/20 | 2281 |
| 12/03/20 | 2876 |
| 13/03/20 | 3661 |
| 14/03/20 | 4499 |
| 15/03/20 | 5423 |
| 16/03/20 | 6633 |
| 17/03/20 | 7730 |
| 18/03/20 | 9134 |
| 19/03/20 | 10995 |
| 20/03/20 | 12612 |
| 21/03/20 | 14459 |
| 22/03/20 | 16689 |
| 23/03/20 | 19856 |
| 24/03/20 | 22302 |
| 25/03/20 | 25233 |
| 26/03/20 | 29155 |
| 27/03/20 | 32964 |
| 28/03/20 | 37575 |
| 29/03/20 | 40174 |
| 30/03/20 | 44550 |
| 31/03/20 | 52128 |
| 01/04/20 | 56989 |
| 02/04/20 | 59105 |
| 03/04/20 | 64338 |
| 04/04/20 | 68605 |

**Supplemental online Table 6**. COVID-19 cumulative incidence by date in Germany.

| DATE | TOTAL CASES |
|---|---|
| 24/02/20 | 16 |
| 25/02/20 | 18 |
| 26/02/20 | 21 |
| 27/02/20 | 26 |
| 28/02/20 | 53 |
| 29/02/20 | 66 |
| 01/03/20 | 117 |
| 02/03/20 | 150 |
| 03/03/20 | 188 |
| 04/03/20 | 240 |
| 05/03/20 | 349 |
| 06/03/20 | 534 |
| 07/03/20 | 684 |
| 08/03/20 | 847 |
| 09/03/20 | 1112 |
| 10/03/20 | 1460 |
| 11/03/20 | 1884 |
| 12/03/20 | 2369 |
| 13/03/20 | 3062 |
| 14/03/20 | 3795 |
| 15/03/20 | 4838 |
| 16/03/20 | 6012 |
| 17/03/20 | 7156 |
| 18/03/20 | 8198 |
| 19/03/20 | 10999 |
| 20/03/20 | 13957 |
| 21/03/20 | 16662 |
| 22/03/20 | 18610 |
| 23/03/20 | 22672 |
| 24/03/20 | 27436 |
| 25/03/20 | 31554 |
| 26/03/20 | 36508 |
| 27/03/20 | 42288 |
| 28/03/20 | 48582 |
| 29/03/20 | 52547 |
| 30/03/20 | 57298 |
| 31/03/20 | 61913 |
| 01/04/20 | 67366 |
| 02/04/20 | 73522 |

**Supplemental online Table 7**. COVID-19 cumulative incidence by date in UK.

| DATE | TOTAL CASES |
|---|---|
| 01/02/20 | 2 |
| 02/02/20 | 2 |
| 03/02/20 | 2 |
| 04/02/20 | 2 |
| 05/02/20 | 2 |
| 06/02/20 | 3 |
| 07/02/20 | 3 |
| 08/02/20 | 3 |
| 09/02/20 | 4 |
| 10/02/20 | 8 |
| 11/02/20 | 8 |
| 12/02/20 | 8 |
| 13/02/20 | 9 |
| 14/02/20 | 9 |
| 15/02/20 | 9 |
| 16/02/20 | 9 |
| 17/02/20 | 9 |
| 18/02/20 | 9 |
| 19/02/20 | 9 |
| 20/02/20 | 9 |
| 21/02/20 | 9 |
| 22/02/20 | 9 |
| 23/02/20 | 9 |
| 24/02/20 | 13 |
| 25/02/20 | 13 |
| 26/02/20 | 13 |
| 27/02/20 | 13 |
| 28/02/20 | 19 |
| 29/02/20 | 23 |
| 01/03/20 | 35 |
| 02/03/20 | 40 |
| 03/03/20 | 51 |
| 04/03/20 | 85 |
| 05/03/20 | 114 |
| 06/03/20 | 160 |
| 07/03/20 | 206 |
| 08/03/20 | 271 |
| 09/03/20 | 321 |
| 10/03/20 | 373 |
| 11/03/20 | 456 |
| 12/03/20 | 590 |
| 13/03/20 | 797 |
| 14/03/20 | 1061 |
| 15/03/20 | 1391 |
| 16/03/20 | 1543 |
| 17/03/20 | 1950 |
| 18/03/20 | 2626 |
| 19/03/20 | 3269 |
| 20/03/20 | 3983 |

| | |
|---|---|
| 21/03/20 | 5018 |
| 22/03/20 | 5683 |
| 23/03/20 | 6650 |
| 24/03/20 | 8077 |
| 25/03/20 | 9529 |
| 26/03/20 | 11658 |
| 27/03/20 | 14548 |
| 28/03/20 | 17104 |
| 29/03/20 | 19606 |
| 30/03/20 | 22271 |
| 31/03/20 | 25521 |
| 01/04/20 | 30088 |
| 02/04/20 | 34610 |
| 03/04/20 | 39282 |
| 04/04/20 | 43282 |
| 05/04/20 | 49481 |
| 06/04/20 | 53624 |
| 07/04/20 | 57512 |
| 08/04/20 | 63377 |
| 09/04/20 | 68052 |
